# Childhood Stress and Amygdala-Hippocampal Volume: A Multiverse Analysis in Adolescents

**DOI:** 10.64898/2026.10.13.26365419

**Authors:** Parnika Sivakumar, Jamie Hanson

## Abstract

Childhood adversity is robustly linked to differences in amygdala and hippocampal volume. However, findings across studies have been inconsistent and contradictory. This inconsistency stems partly from the many reasonable but arbitrary choices that researchers make when operationalizing brain regions, stress exposure, and covariates. To deal with this variability, we applied a multiverse analysis approach using adolescent data from the Boston Adolescent Neuroimaging of Depression and Anxiety (BANDA) study to evaluate how multiple analytic choices influence the relationships between stress and hippocampal/amygdala volume. Our primary analysis crossed three region-of-interest operationalizations (left, right, total), four intracranial volume scaling methods, and multiple sociodemographic covariates. Greater lifetime stress severity was associated with smaller amygdala volume across 72% of specifications, and with smaller hippocampal volume across 52% of specification models. Statistical significance was strongly influenced by the region of interest. Our secondary multiverse analysis tested six operationalizations of stress exposure (total/acute/chronic x count/severity). This analysis found that the laterality pattern in the primary analysis was not strongly influenced by stress operationalization. These findings demonstrate the robustness of the stress-medial temporal lobe association and identify specific analytic choices (i.e. hemisphere operationalization) that most strongly drive the variability observed in previous literature. Our study is one of the first that utilized multiverse analysis for adversity-neuroimaging research, and provides a strong framework for evaluating and interpreting inconsistent findings.

## INTRODUCTION

Childhood adversity can be detrimental to an individual’s physical, emotional, and social development. Experiences such as physical abuse and neglect have been associated with higher rates of substance use, psychopathology, and physical health issues (Madigan et al., 2023; McLaughlin et al., 2019). Despite these findings being well-replicated in the literature, the *pathways* connecting childhood adversity and its multidimensional negative outcomes are still poorly understood.

To make progress in this space, research has turned to neurobiology, focusing on medial temporal brain regions like the hippocampus and amygdala. The hippocampus guides the formation of declarative and contextual memory and plays a part in stress responsivity (Kim & Diamond, 2002). The amygdala evaluates incoming emotional experiences, and is critical for fear learning, aggression, and stress regulation (Roozendaal et al., 2009; Rosell & Siever, 2015). Research has generally suggested smaller volumes and differences in functional connectivity in both brain regions post-adversity (Gorka et al., 2014; Hanson et al., 2015, 2019; Kahhalé et al., 2023; Tottenham & Sheridan, 2010). However, volumetric MRI results have been variable and inconsistent depending on the type (abuse vs. neglect) and timing of the adversity, with differences often found in the direction and magnitude of reported effects (McLaughlin et al., 2019).

Inconsistencies in adversity research could be due to many factors, spanning conceptual and methodological elements of different studies. Conceptually, how adversity is modeled and when it is examined during development could be contributing to inconsistent findings (Hanson & Nacewicz, 2021; McLaughlin et al., 2021; Miller et al., 2018). Methodologically, variability in study design and analytic choice could be creating some substantial differences in the results and decreasing reproducibility (McLaughlin et al., 2019; Nosek et al., 2022). Given the striking prevalence of childhood adversity and the potential impact of neurobiological differences on overall health, establishing more replicable findings in this area is essential.

To deal with variability in analytic choices, researchers are increasingly turning to “multiverse analysis”, an approach where different combinations of analytic choices create a ‘multiverse’ of possible results (Steegen et al., 2016). Most commonly, a single analysis is conducted on a single dataset based on a *single* choice about the combination, transformation, and categorization of data variables. In a multiverse analysis, we conduct multiple analyses to map how sensitive the final conclusions are to a variety of analytic choices. Prominent examples in social science research have underscored that results can vary substantially depending on how key variables are operationalized and what covariates are included (Steegen et al., 2016). However, there is limited application of multiverse analysis to neuroimaging research (cf. (Bloom et al., 2022). The application of such an approach to neuroimaging research can showcase how different analytic decisions influence the robustness and replicability of neuroimaging findings.

As such, multiverse analysis could be particularly valuable, especially with regard to childhood adversity and neurodevelopment. Here, we took a multiverse approach to examine the relationship between early life adversity and both amygdala and hippocampal volume, using a range of analytic choices that are known sources of inconsistency in neuroimaging research. We made decisions regarding right, left, or total hemispheric volume, whole-brain scaling methods, and the inclusion or exclusion of different covariates (e.g., age, sex, race). Using a publicly-available MRI dataset with robust measures of stress and adversity, we hypothesized that greater early life adversity would be significantly associated with smaller volumes in both the amygdala and the hippocampus. We anticipated that these associations would remain significant across the majority of the analytic specifications within our multiverse. We also explored whether the exact operationalization of adversity led to variations in results in a secondary analysis. By comprehensively examining analytic variations, we hope to clarify past discrepancies and report the most stable and reproducible relations between childhood adversity and medial temporal lobe neurobiology.

## METHODS

### Participants

This study analyzed data from the Boston Adolescent Neuroimaging of Depression and Anxiety (BANDA) study. The study included 215 adolescents aged 14-17. All participants completed structural, diffusion, and functional MRI scans, along with clinical and behavioral questionnaires over four sessions. Twelve participants were excluded due to unusable or missing structural MRI data, yielding a final analytic sample of 203.

**Table 1.** Sample Demographic Characteristics.

| Characteristic | N = 203<sup>1</sup> |
| --- | --- |
| **Age, years** | 15.42 (14.75, 16.00) |
| **Sex** |  |
| Female | 126 (64%) |
| Male | 71 (36%) |
| **Race** |  |
| White | 150 (79%) |
| Non-White | 40 (21%) |
| **Ethnicity** |  |
| Not Hispanic/Latino | 183 (93%) |
| Hispanic/Latino | 14 (7.1%) |
| **Parental Education** |  |
| 4-year or higher | 166 (85%) |
| Some college | 22 (11%) |
| ≤ High School/Vocational | 8 (4.1%) |
| **Pubertal Stage (Tanner Scale)** | 3.60 (3.20, 3.80) |
| <sup>1</sup>Median (Q1, Q3); n(%). Percentages calculated among non-missing responses; missing data ranged from n=6 (Sex, Ethnicity) to n=13 (Race) across characteristics. |  |

### Neuroimaging Acquisition and Preprocessing

T1-weighted, high resolution structural images were obtained via a 3T Siemens Prisma scanner and a 64-channel head coil. Image files were assessed by visual inspection for artifacts that could interfere with analyses (Siless et al., 2020). Structural images first underwent HCP minimal preprocessing (Glasser et al., 2013), which included gradient distortion correction, alignment and averaging of T1 and T2-weighted volumes, bias-field correction for intensity normalization, and nonlinear registration to MNI152 standard space. Images were then run through FreeSurfer’s (version 7.1) recon-all pipeline, which generated initial white-matter and pial surface boundaries. Subcortical volume segmentation was performed using the Freesurfer automated subcortical segmentation pipeline (aseg), which labels neuroanatomical structures based on a set of manually segmented reference scans (Fischl et al., 2002). Through this pipeline, we obtained amygdala and hippocampal volumetric estimates (left, right, and total) (Kahhale et al., 2023). Whole-brain volume measures (e.g., total gray matter volume, and estimated total intracranial volume [eTIV]), used for ICV scaling, were also obtained via this processing pipeline.

### Stress Exposure

Stress was measured using the Stress and Adversity Inventory for Adolescents (Adolescent STRAIN). The STRAIN is an online survey that assesses lifetime exposure to 75 different stressors, 33 of which are acute (short, episodic experiences) and 42 of which are chronic (long-term, across the lifespan). For each stressor, respondents answer follow-up questions about the stressor’s severity, frequency, timing, and duration. Stress exposure summary scores are then calculated to summarize the total lifetime stressor count and severity, across each stressor and across life domains, making for a uniquely cohesive and detailed overview of stress exposure in individuals (Slavich et al., 2019). This inventory addresses the limitation of treating adversity as a singular construct by capturing both cumulative and micro-domain stress exposure across twelve life domains (e.g., housing, marital/partner relationships, education, etc.) and five social-psychological characteristics (e.g., Interpersonal Loss, Physical Danger, Humiliation, Entrapment, Role Change), with higher scores indicating greater stress exposure (Slavich et al., 2019). The STRAIN has demonstrated strong predictive validity, showing significant associations with a range of mental and physical health outcomes, including mood disorders, risky behaviors, and clinical diagnoses (Slavich et al., 2019).

In our primary analysis, we operationalized stress using the STRAIN’s Total Lifetime Stressor Severity Score. This summary score sums up severity ratings across all 75 different stressors into a single, continuous index of cumulative lifetime adversity, with higher scores indicating greater stress severity. The STRAIN also generates a Total Lifetime Stressor Count Score, as well as severity and count scores specific to acute and chronic stressors. We however limited our representation of stress in our primary analysis to the STRAIN’s Severity Score, while varying different covariates (e.g. age & sex, race, parental education, puberty, CAT12).

### Decision Points and Covariates

Consistent with our multiverse approach, we identified two structural decision points. The first was which regions of interest to include. Rather than solely looking at total amygdala and hippocampal volume, we observed total, right, and left volume, as left and right volumes may respond differently to stress.

In our primary analysis, our second decision point was how brain volume measures were corrected. Generally, brain volume measures are corrected for individual differences in head size, but studies vary in how this correction is performed. To account for this variability, we included four different approaches to intracranial volume scaling: no correction, total brain segmentation, total gray matter volume, and estimated total intracranial volumes (eTIV).

Our secondary analysis differed in its second decision point. Here we explored the operationalization of stress rather than intracranial volume measures, as there is variability in how stress can be defined, with there being acute versus chronic stress measurements, as well as stressor count versus stressor severity. The STRAIN survey accounts for this variety of stress definitions, and we decided to delve further to test whether our primary analysis findings depended on the specific operationalization of stress. We conducted a secondary multiverse analysis with six alternative STRAIN summary scores: acute severity, acute count, chronic severity, chronic count, total severity, and total count. Through this secondary analysis, we aimed to assess whether stress associations previously observed with the amygdala and hippocampus reflect a general relationship with cumulative adversity, or are specific to how “stress” is measured (total/acute/chronic x count/severity).

Along with the two forking paths (intracranial volume scaling and region of interest operationalization), part of multiverse analysis is making decisions about which covariates to include, as including or excluding a given covariate could alter the observed association between brain volume and stress. We treated all covariates as binary variables (present vs. absent) in each model specification. Our primary analysis used the following five covariates: age & sex, race, parental education, pubertal status, and CAT12 grading. Age & sex was included or excluded together in their original coding respectively, and race was included as a binary variable. Parental education was recoded into a 3-level factor: 1= “≤ HS/Vocational”, 2 = “Some college”, and 3 = “4-yr or higher”. Pubertal status was measured through the Tanner Scale, a 5-point rating scale for physical development status (including primary and secondary sex characteristics), ranging from pre-pubescence through adulthood. Computational Anatomy Toolbox Version 12 (CAT12), a scan image quality grade used for morphometric analyses, was originally coded as a continuous variable. Past work has found relations between these covariates and medial temporal lobe volumes (Gilmore et al., 2021; Hanson et al., 2011, 2026).

Our secondary analysis held the following covariates constant rather than treating them as binary variables: age, sex, pubertal status, and eTIV (estimated total intracranial volume). We decided to keep a minimal number of covariates and use eTIV as a covariate rather than as a forked decision point in order to lessen the number of models run and more clearly observe the stress operationalization variability on results.

### Analysis Plan

In our primary analysis, we ran linear regression models with amygdala or hippocampal volume (left, right, or total) as the independent variable. Total Lifetime Stressor Severity was the dependent variable in this analysis. The two structural forks (ROI operationalization x ICV scaling) and the five covariate forks yield 3 × 4 × 2⁵ = 384 unique model specifications per brain region (see Figure 1). Together, these specifications allow us to assess both robustness and sensitivity by demonstrating whether results hold up regardless of analytic choices and which decision points drive the most variability in our results. Each model was ranked by statistical significance (p < .05) and then by absolute effect size within each significance band.

**Figure 1.**
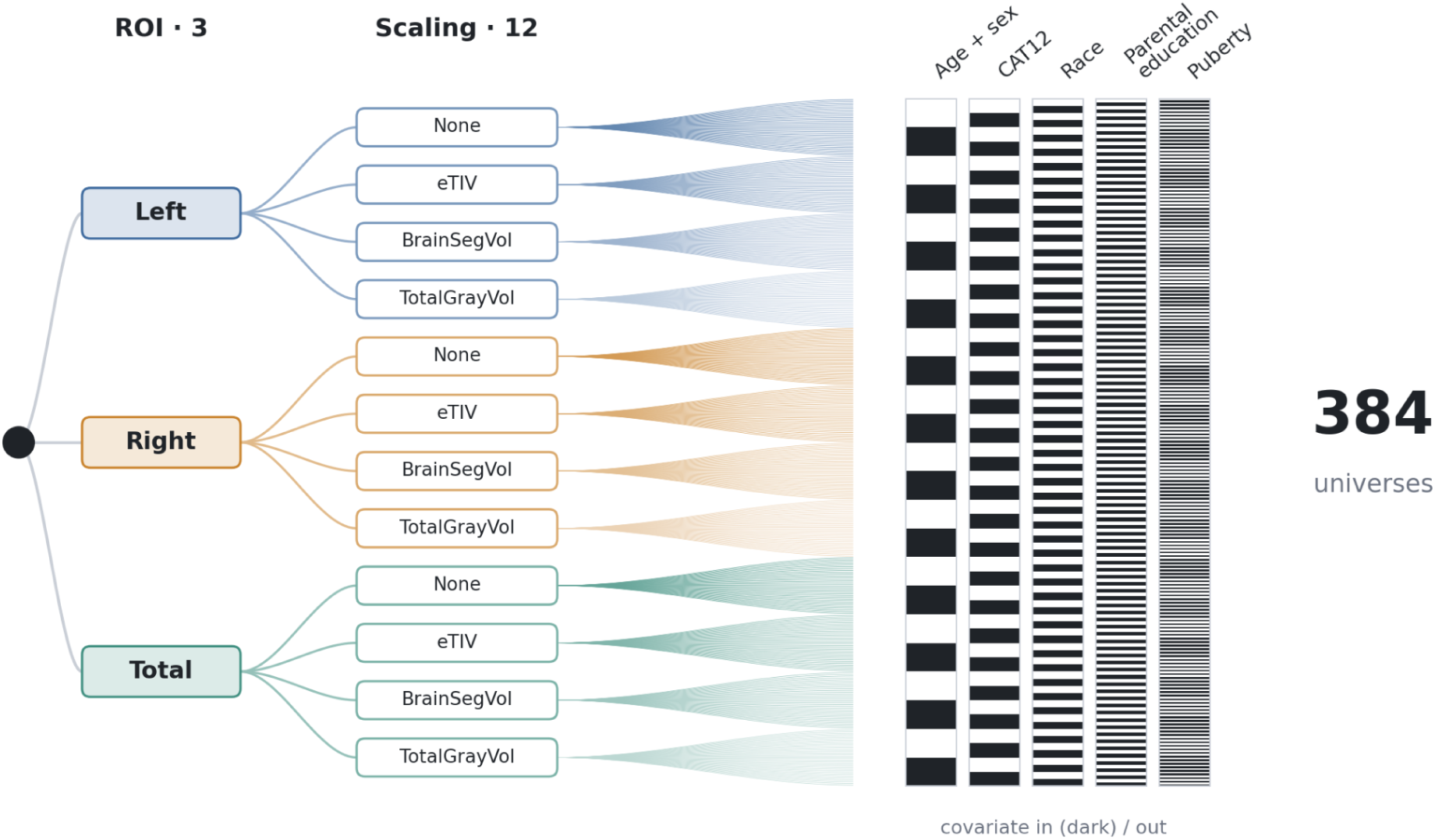
Primary Analysis: Multiverse Forking Paths The figure above displays the two structural forks in multiverse analysis: (1) how the region of interest (ROI) is operationalized (left, right or total volume), and (2) how that volume is scaled for individual differences in head/brain size (none, estimated total intracranial volume - “eTIV”, total brain segmentation volume - “BrainSegVol”, or total gray matter volume - “TotalGrayVol”). Crossing these two forks produces 3 ROI x 4 ICV = 12 unique structural specifications for the amygdala and hippocampus brain regions respectively. With five additional covariates (age & sex, CAT12, race, parental education, and pubertal status) used as binary variables in our analysis, we have 2^5 possible covariate combinations. In total, that gives us 3 x 4 x 2^5 = 384 unique model specifications/universes for the amygdala and hippocampus brain regions respectively.

Our secondary analysis was run similarly, as linear regression models with amygdala or hippocampal volume (left, right, or total) as the independent variable. The dependent variable, stress, was operationalized in six different ways: total, chronic, or acute, each scored as a count and severity rating. This allowed us to observe the association between specific stressor types/scores and a corresponding hippocampal or amygdala region, and whether or not these associations maintain robustness across the six stress operationalizations. The two structural forks in this analysis (region of interest x stress) and four fixed covariates (age, sex, pubertal status, and eTIV) yielded 6 x 3 = 18 different model specifications per brain region (see Figure 2). Each model was ranked by statistical significance (p < .05) and then by absolute effect size within each significance band.

**Figure 2.**
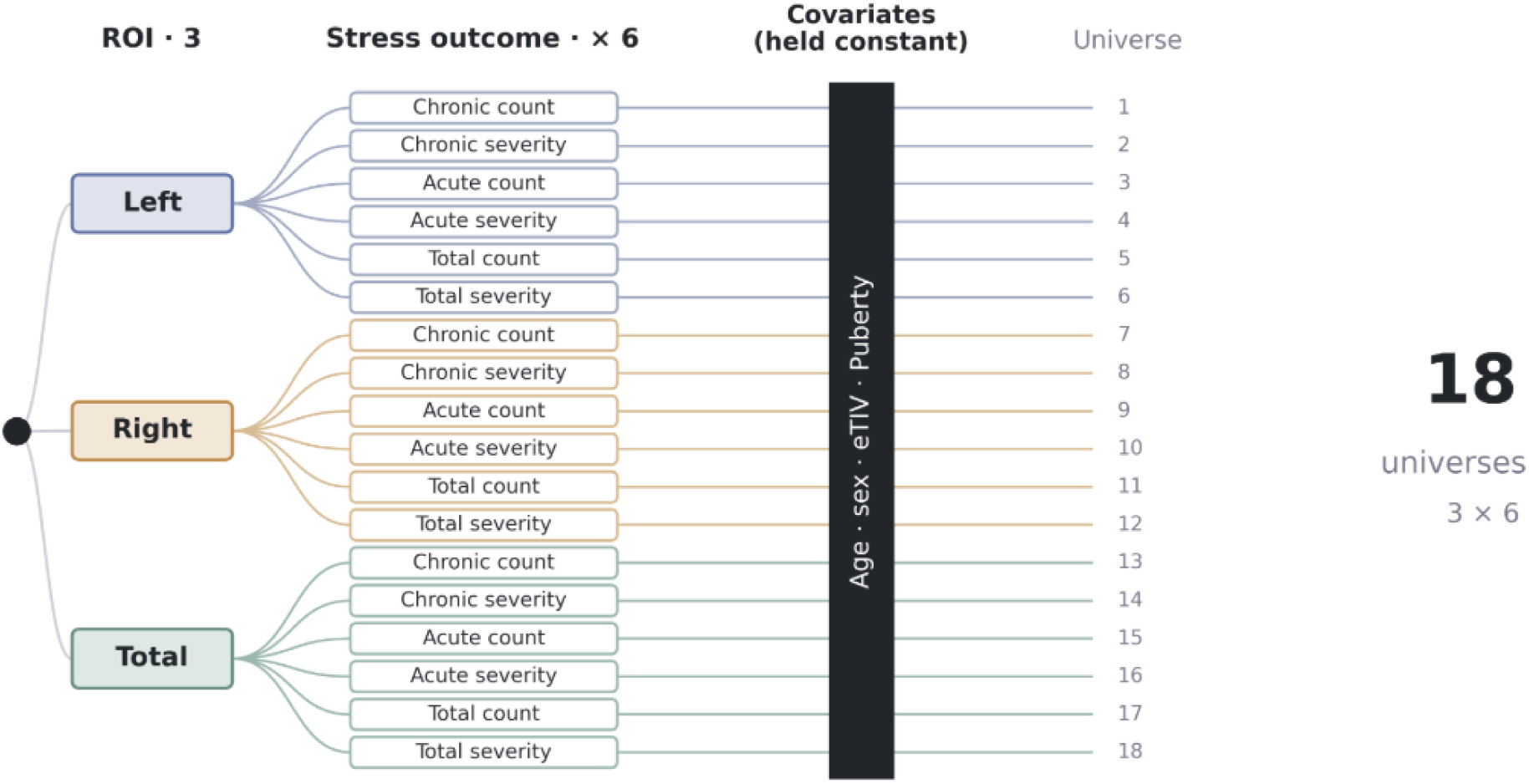
Secondary Analysis: Multiverse Forking Paths The figure above displays the two structural forks in this secondary multiverse analysis: (1) how the region of interest (ROI) is operationalized (left, right or total volume), and (2) how the “stress” outcome is defined (chronic vs. acute and count vs. severity). Crossing these two forks produces 3 ROI x 6 stress operationalizations = 18 unique structural specifications for the amygdala and hippocampus brain regions respectively. Covariates (age, sex, eTIV, and puberty) were kept constant in this secondary analysis.

## RESULTS

### Primary Analysis

Across 384 model specifications, amygdala and hippocampal volumes were negatively associated with stress severity (all point estimates from each specification model were negative). 277 out of 384 amygdala models and 198 out of 384 hippocampus models were statistically significant (p<.05). Specification curves are shown in Figures 3 and 4. Full model-by-model results (β, SE, t, p, 95% CI) for all 384 specifications are provided in Supplementary Tables S1 (hippocampus) and S2 (amygdala).

**Figure 3.**
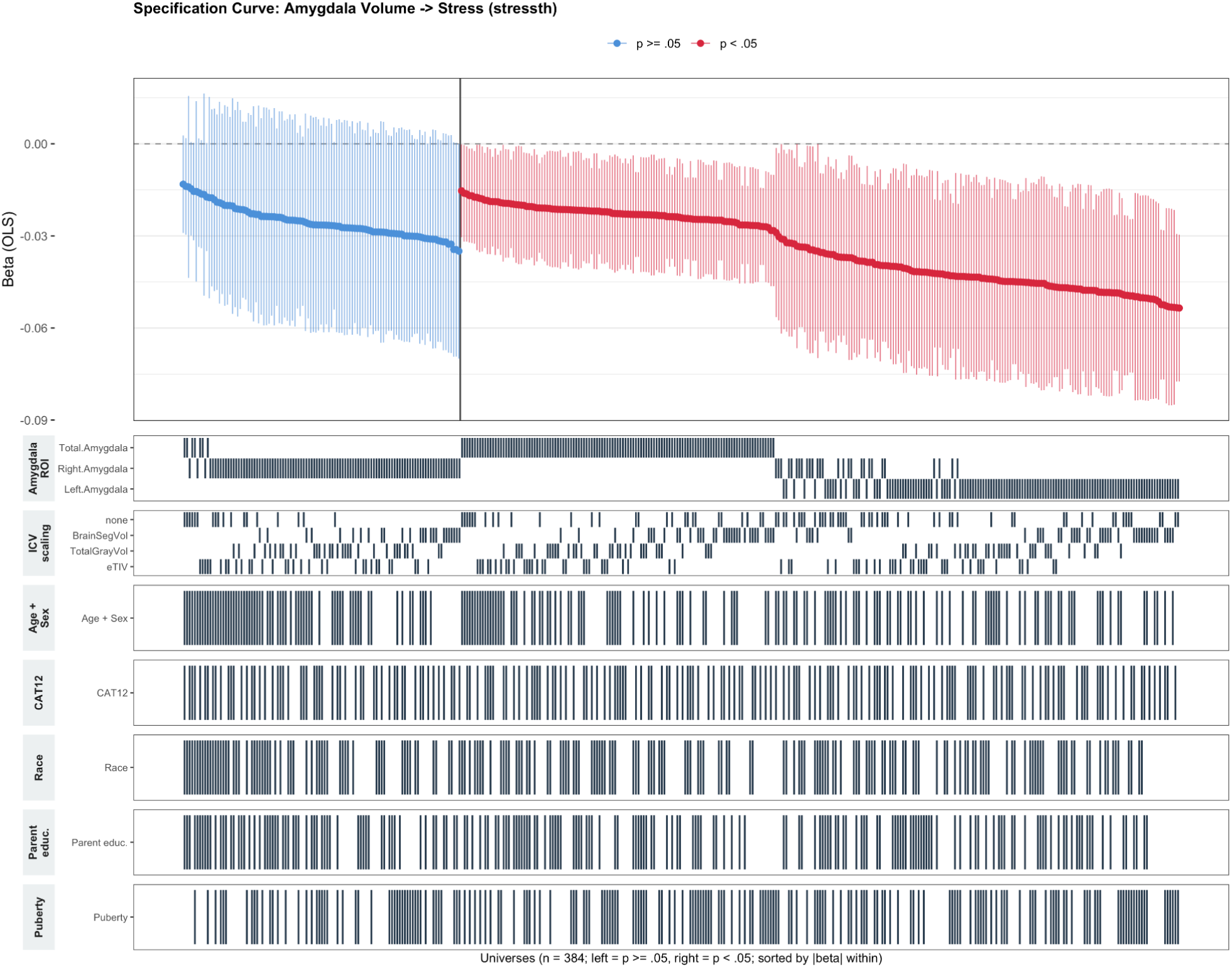
Primary Analysis: Specification Curve – Amygdala This specification curve displays the 384 different model specifications for amygdala volume, with outcome stress. Each point represents the estimated beta coefficient for a single model specification (hence 384 lines total). All primary estimates are unstandardized. Vertical lines indicate 95% confidence intervals, and specifications are ordered by effect size within each significance group. Blue points/lines indicate insignificant findings (p>0.05), while red points/lines indicate significant findings (p<0.05). The tile matrix below the plot shows which analytic choices were used for each specification (e.g., amygdala operationalization, ICV scaling, and covariate inclusion), aligned with the corresponding point above. Clusters within a given row of the tile matrix - for example, left- and total-amygdala specifications clustering almost entirely on the significant (red) side, while right-amygdala specifications cluster on the insignificant (blue) side - indicates that this analytic choice is a strong driver of whether the association reaches statistical significance.

**Figure 4.**
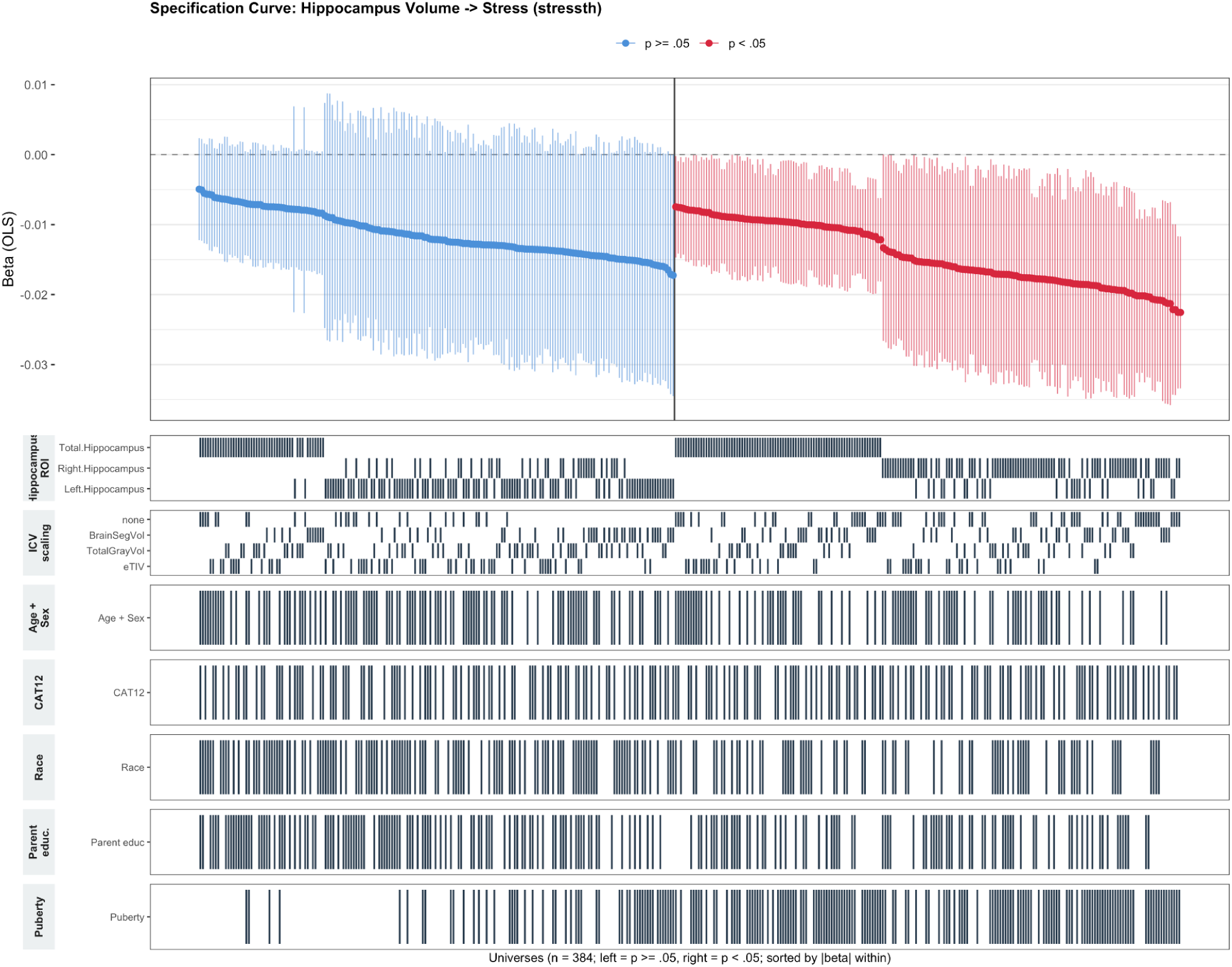
Primary Analysis: Specification Curve – Hippocampus This specification curve displays the 384 different model specifications for hippocampal volume, with outcome stress. Each point represents the estimated beta coefficient for a single model specification (hence 384 lines total). Vertical lines indicate 95% confidence intervals, and specifications are ordered by effect size within each significance group. Blue points/lines indicate insignificant findings (p>0.05), while red points/lines indicate significant findings (p<0.05). The tile matrix below the plot shows which analytic choices were used for each specification (e.g., hippocampus operationalization, ICV scaling, and covariate inclusion), aligned with the corresponding point above. Right-hippocampus specifications clustering almost entirely on the significant (red) side, while left-hippocampus specifications cluster on the insignificant (blue) side, indicate the reverse of the laterality pattern observed in the amygdala analysis, where left-hemisphere volume was the more consistently significant operationalization.

Significance was strongly driven by the region of interest. For example, the left amygdala volume was significant in 100% of specifications, the total volume significant in 94.5% of specifications, and the right amygdala volume significant in 21.9% of specifications. In the hippocampus, we notice the inverse: right hippocampal volume was significant in 69.5% of specifications, total hippocampal volume was significant in 63.3% of specifications, and left hippocampal volume was significant in only 21.9% of specifications. This pattern indicates that the main source of specification-dependent variability in this multiverse analysis was that of brain region operationalization.

### Secondary Analysis

Our analysis confirms that brain-stress associations in the amygdala and hippocampus are generalizable across the six different operational definitions of stress exposure. In the amygdala, 15 out of 18 specifications (83.3%) were significant, with left-and total-amygdala specifications being significant in 100% of cases, and right-amygdala specifications being significant in only 50% (see Figure 5). In the hippocampus, 14 out of 18 specifications (77.8%) were significant, with right-hippocampus specifications significant in 100% of cases, total-hippocampus specifications significant in 83.3% of cases, and left-hippocampus specifications significant in only 50% of cases (see Figure 6). Full model-by-model results for all 18 specifications per region are provided in Supplementary Tables S3 (hippocampus) and S4 (amygdala).

**Figure 5.**
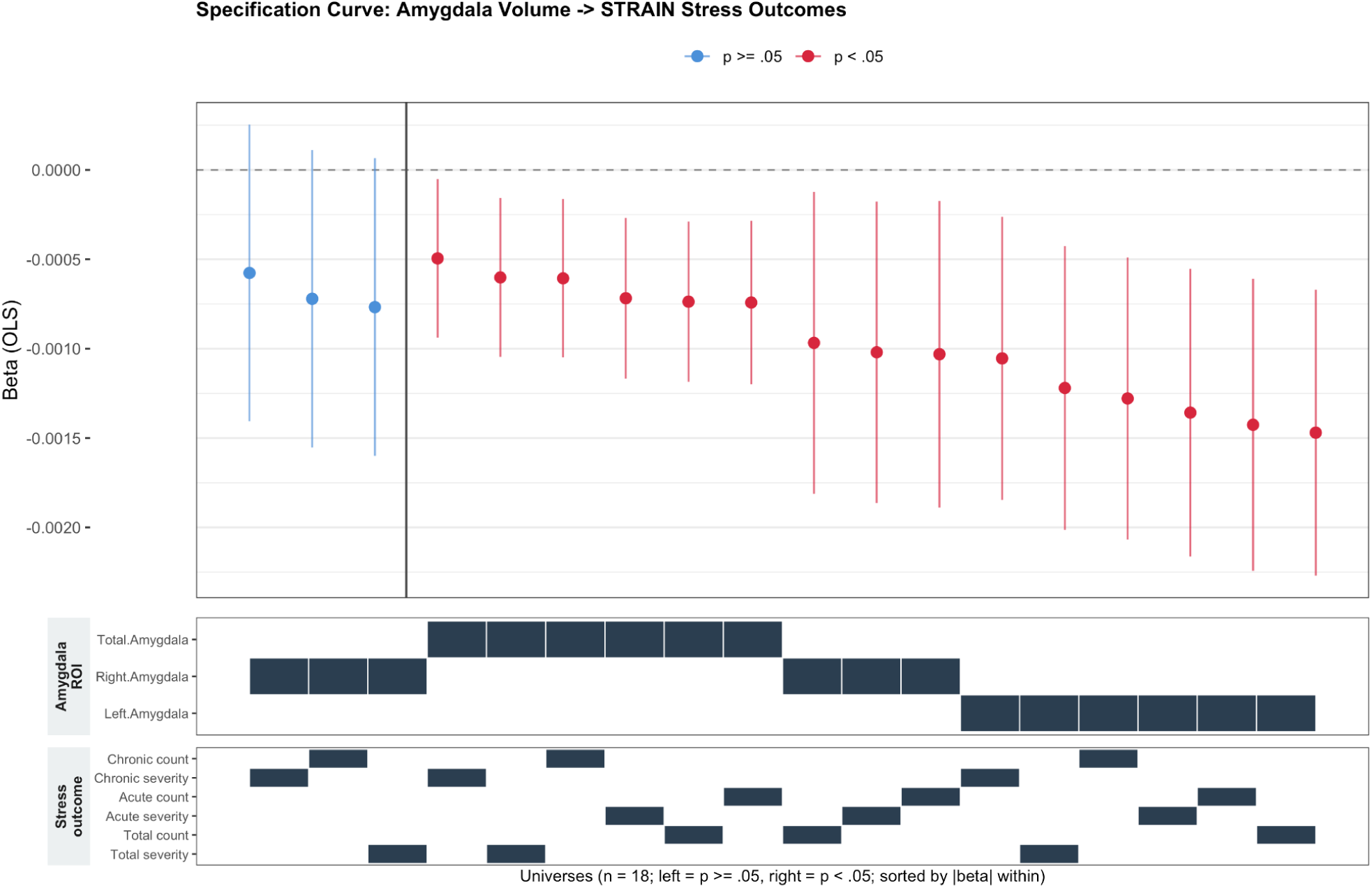
Secondary Analysis: Specification Curve - Amygdala The figure above shows standardized beta coefficients (points) and 95% confidence intervals (vertical lines) for the association between amygdala volume and six alternative STRAIN stress outcome variables (acute, chronic, and total stressor count and severity), crossed with amygdala specification (left, right, total), yielding 18 model specifications. Specifications are ordered by effect size within each significance group. Blue points/lines represent insignificant findings (p>0.05), while red points/lines represent significant findings (p<0.05). The tile matrix (bottom) indicates amygdala ROI and stress outcome variable operationalization used in each model specification.

**Figure 6.**
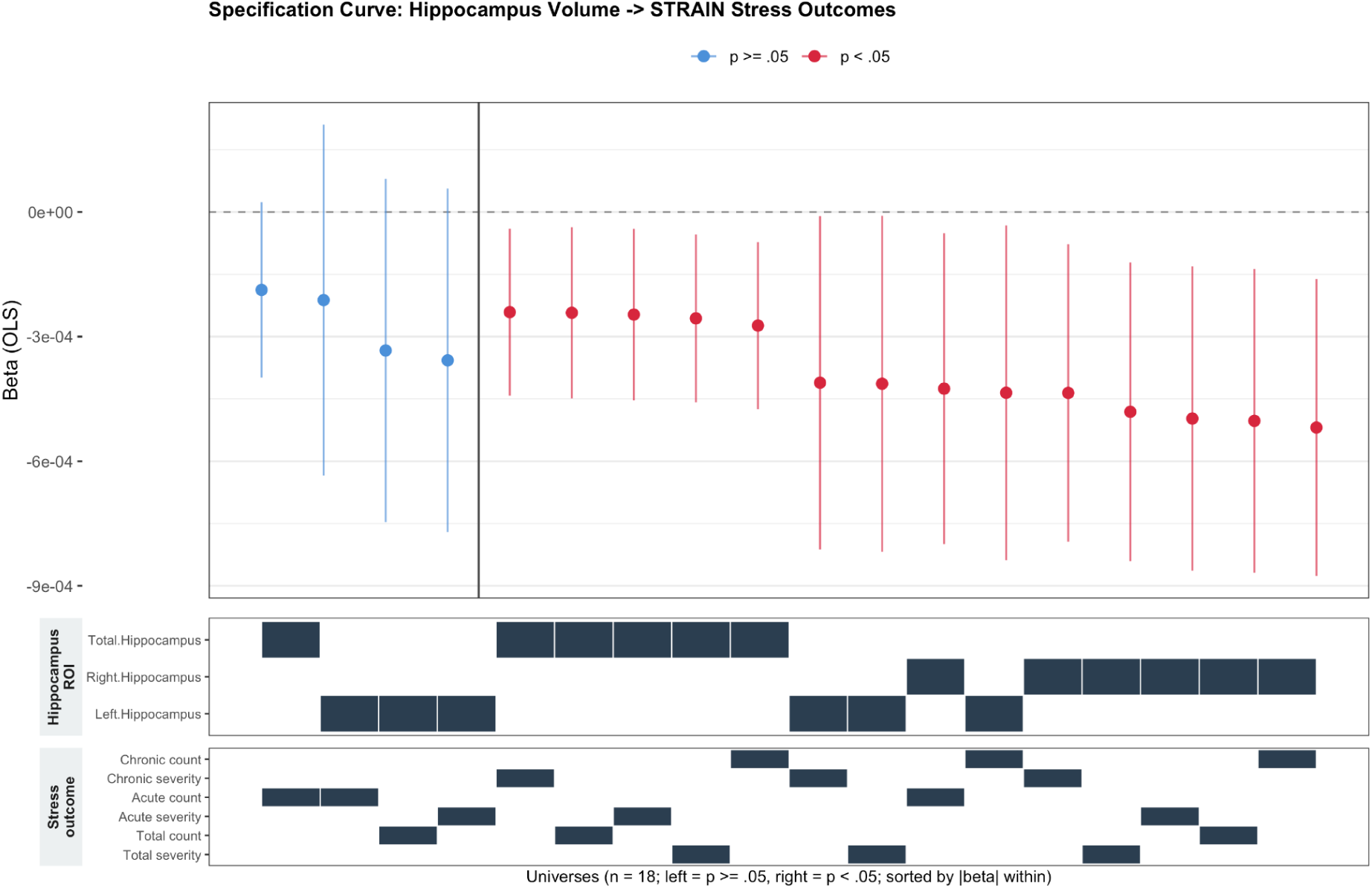
Secondary Analysis: Specification Curve – Hippocampus The figure above shows standardized beta coefficients (points) and 95% confidence intervals (vertical lines) for the association between hippocampal volume and six alternative STRAIN stress outcome variables (acute, chronic, and total stressor count and severity), crossed with hippocampus ROI specification (left, right, total), yielding 18 model specifications. Specifications are ordered by effect size within each significance group. Blue points/lines represent insignificant findings (p>0.05), while red points/lines represent significant findings (p<0.05). The tile matrix (bottom) indicates hippocampus ROI and stress outcome variable operationalization used in each model specification.

These results mirror that of our primary analysis, indicating that the laterality pattern stays consistent across all operationalization of stress exposure.

## DISCUSSION

This study used a multiverse analysis approach to explore the association between stress exposure and amygdala and hippocampal volume among adolescents. Our primary analysis assessed this relationship across 384 model specifications, varying region-of-interest (ROI) operationalization, intracranial volume (ICV) scaling method, and covariate inclusion. Overall, greater stress severity was associated with smaller amygdala and hippocampal volumes. The strength of the association was driven primarily by how the ROI was operationalized, with left and total amygdala volume, and right hippocampal volume consistently achieving significant results. A secondary analysis tested whether these findings depended on the operationalization of stress (total/acute/chronic x count/severity). How stress was defined had little influence on the overall pattern of results, indicating that it did not drive the strength of the association.

The findings of this analysis are generally consistent with prior work linking greater early life stress to smaller amygdala and hippocampal volume (Hanson et al., 2015; McLaughlin et al., 2019)). The contribution of this study lies less in the direction of the association, and more in demonstrating how robust that association is with the accounting of a wide range of analytic choices. Single-specification analyses report a result under one specific set of analytic decisions, and often note that alternative operationalizations were not accounted for as a part of their study limitations. By systematically varying operationalization of ROI, ICV, stress, and covariate inclusion, this study provides a more complete picture of which analytic choices are meaningful, and which choices are not. This analysis furthers the application of multiverse analysis to adversity-neuroimaging research (Bloom et al., 2022) and provides a framework for future research in this area to assess the stability of findings against unexplored analytic variability.

While this multiverse analysis addresses a key limitation of prior single-specification studies, it is not without its own limitations. First, no multiverse analysis is truly exhaustive. We controlled (1) which analytic decisions to treat as forks and (2) which levels/options to include within each fork. There are many other reasonable analytic choices that could have further influenced the observed results (i.e. alternative brain segmentation approaches, volumetric estimates). The forks included here were selected based on documented sources of variability in previous literature, but this is a bounded rather than a complete decision space. Second, despite accounting for race and parental education as covariates, selection bias may be present in the underlying BANDA sample. Because BANDA samples include a substantial portion of participants with current anxiety/depression diagnoses, and more anxious individuals tend to move more in the scanner, it is possible that participants with more severe symptomatology, who may also report higher stress exposure, were disproportionately excluded due to unusable structural scans. This exclusion could disproportionately remove high-stress cases from the analytic sample. Finally, stress exposure was measured retrospectively via self-report using the STRAIN, which may be subject to recall bias. Therefore, the accuracy of reporting stressor timing, frequency, and severity cannot be verified.

Our current findings support a robust directional association between early life stress and reduced amygdala and hippocampal volume in adolescence, with statistical significance depending heavily on hemispheric region. This study demonstrates the value of using multiverse analysis as a tool for assessing the stability of neuroimaging findings, and would be useful for future investigation.

## Data Availability

All data produced are available online at https://www.humanconnectome.org/study/connectomes-related-anxiety-depression/data-releases.

## Declaration of the use of AI

AI was used to check spelling and grammar. There was also use to reword small portions of draft text (2-5%) and reformatting of supplemental materials. These operations were completed using Claude Sonnet 5.

## Data and Code Availability

Data were provided by the Boston Adolescent Neuroimaging of Depression and Anxiety (BANDA) Consortium’s Human Connectome Project, supported by U01MH108168 (PIs: Drs. Susan Whitfield-Gabrieli and John Gabrieli).

## Author Contributions

Ms. Sivakumar created tables and figures; and co-authored the manuscript. Dr. Hanson processed the data and generated analysis pipelines; created figures; and co-authored the manuscript.

## Supplementary Material

Supplemental tables for all analytical combinations are provided. https://docs.google.com/document/d/1oPChiw04vnnojHdaCmWdrJmluV8DeVmZ3wzAtLfx5Hk/edit?usp=sharing

