## Supplemental Material for "Childhood Stress and Amygdala-Hippocampal Volume: A Multiverse Analysis in Adolescents"

**Table S1. Multiverse results: hippocampus volume → stress severity (stressth)**

*All 384 model specifications (3 ROI operationalizations × 4 ICV scaling methods × 5 binary covariates). β = unstandardized OLS coefficient; SE = standard error; CI = 95% confidence interval. Values rounded to four decimal places.*

| **model_label** | **estimate** | **SE** | **t** | **p** | **CI low** | **CI high** | **significant** |
| --- | --- | --- | --- | --- | --- | --- | --- |
| Total.Hippocampus \| age+sex+CAT12+race+parent_educ | -0.0050 | 0.0037 | -1.3360 | 0.1833 | -0.0122 | 0.0024 | FALSE |
| Total.Hippocampus \| age+sex+race+parent_educ | -0.0050 | 0.0037 | -1.3658 | 0.1737 | -0.0123 | 0.0022 | FALSE |
| Total.Hippocampus \| age+sex+CAT12+race | -0.0055 | 0.0036 | -1.5315 | 0.1274 | -0.0127 | 0.0016 | FALSE |
| Total.Hippocampus \| age+sex+race | -0.0057 | 0.0036 | -1.5679 | 0.1187 | -0.0128 | 0.0015 | FALSE |
| Total.Hippocampus \| age+sex+EstimatedTotalIntraCranialVol+race+parent_educ | -0.0057 | 0.0041 | -1.3990 | 0.1636 | -0.0137 | 0.0023 | FALSE |
| Total.Hippocampus \| age+sex+EstimatedTotalIntraCranialVol+CAT12+race+parent_educ | -0.0058 | 0.0041 | -1.4215 | 0.1570 | -0.0138 | 0.0022 | FALSE |
| Total.Hippocampus \| age+sex+CAT12+parent_educ | -0.0061 | 0.0036 | -1.7180 | 0.0875 | -0.0132 | 0.0009 | FALSE |
| Total.Hippocampus \| age+sex+parent_educ | -0.0062 | 0.0036 | -1.7391 | 0.0837 | -0.0132 | 0.0008 | FALSE |
| Total.Hippocampus \| age+sex+EstimatedTotalIntraCranialVol+race | -0.0063 | 0.0040 | -1.5668 | 0.1189 | -0.0141 | 0.0016 | FALSE |
| Total.Hippocampus \| age+sex+EstimatedTotalIntraCranialVol+CAT12+race | -0.0063 | 0.0040 | -1.5867 | 0.1144 | -0.0142 | 0.0015 | FALSE |
| Total.Hippocampus \| TotalGrayVol+race+parent_educ | -0.0064 | 0.0045 | -1.4047 | 0.1619 | -0.0153 | 0.0026 | FALSE |
| Total.Hippocampus \| TotalGrayVol+CAT12+race+parent_educ | -0.0064 | 0.0045 | -1.4103 | 0.1602 | -0.0154 | 0.0026 | FALSE |
| Total.Hippocampus \| age+sex+EstimatedTotalIntraCranialVol+parent_educ | -0.0065 | 0.0040 | -1.6467 | 0.1013 | -0.0143 | 0.0013 | FALSE |
| Total.Hippocampus \| EstimatedTotalIntraCranialVol+race+parent_educ | -0.0066 | 0.0041 | -1.6176 | 0.1075 | -0.0147 | 0.0015 | FALSE |
| Total.Hippocampus \| age+sex+EstimatedTotalIntraCranialVol+CAT12+parent_educ | -0.0067 | 0.0040 | -1.6818 | 0.0943 | -0.0145 | 0.0012 | FALSE |
| Total.Hippocampus \| EstimatedTotalIntraCranialVol+CAT12+race+parent_educ | -0.0067 | 0.0041 | -1.6233 | 0.1063 | -0.0148 | 0.0014 | FALSE |
| Total.Hippocampus \| TotalGrayVol+parent_educ | -0.0068 | 0.0044 | -1.5430 | 0.1245 | -0.0154 | 0.0019 | FALSE |
| Total.Hippocampus \| TotalGrayVol+CAT12+parent_educ | -0.0068 | 0.0044 | -1.5549 | 0.1217 | -0.0155 | 0.0018 | FALSE |
| Total.Hippocampus \| age+sex+CAT12+race+parent_educ+puberty | -0.0069 | 0.0038 | -1.8428 | 0.0671 | -0.0144 | 0.0005 | FALSE |
| Total.Hippocampus \| age+sex+race+parent_educ+puberty | -0.0071 | 0.0038 | -1.8783 | 0.0620 | -0.0145 | 0.0004 | FALSE |
| Total.Hippocampus \| EstimatedTotalIntraCranialVol+parent_educ | -0.0071 | 0.0040 | -1.7798 | 0.0768 | -0.0149 | 0.0008 | FALSE |
| Total.Hippocampus \| TotalGrayVol+race | -0.0071 | 0.0045 | -1.5836 | 0.1150 | -0.0160 | 0.0018 | FALSE |
| Total.Hippocampus \| TotalGrayVol+CAT12+race | -0.0072 | 0.0045 | -1.5858 | 0.1145 | -0.0160 | 0.0017 | FALSE |
| Total.Hippocampus \| age+sex+TotalGrayVol+race+parent_educ | -0.0072 | 0.0045 | -1.6066 | 0.1100 | -0.0159 | 0.0016 | FALSE |
| Total.Hippocampus \| EstimatedTotalIntraCranialVol+CAT12+parent_educ | -0.0072 | 0.0040 | -1.7915 | 0.0749 | -0.0150 | 0.0007 | FALSE |
| Total.Hippocampus \| age+sex+TotalGrayVol+CAT12+race+parent_educ | -0.0074 | 0.0045 | -1.6515 | 0.1004 | -0.0162 | 0.0014 | FALSE |
| Total.Hippocampus \| age+sex+BrainSegVol+race+parent_educ | -0.0074 | 0.0044 | -1.6862 | 0.0935 | -0.0161 | 0.0013 | FALSE |
| Total.Hippocampus \| age+sex+EstimatedTotalIntraCranialVol+race+parent_educ+puberty | -0.0075 | 0.0041 | -1.8237 | 0.0699 | -0.0155 | 0.0006 | FALSE |
| Total.Hippocampus \| EstimatedTotalIntraCranialVol+race | -0.0075 | 0.0040 | -1.8454 | 0.0666 | -0.0154 | 0.0005 | FALSE |
| Total.Hippocampus \| BrainSegVol+race+parent_educ | -0.0075 | 0.0045 | -1.6586 | 0.0990 | -0.0163 | 0.0014 | FALSE |
| Total.Hippocampus \| EstimatedTotalIntraCranialVol+CAT12+race | -0.0075 | 0.0041 | -1.8466 | 0.0665 | -0.0155 | 0.0005 | FALSE |
| Total.Hippocampus \| age+sex+EstimatedTotalIntraCranialVol+CAT12+race+parent_educ+puberty | -0.0075 | 0.0041 | -1.8330 | 0.0685 | -0.0156 | 0.0006 | FALSE |
| Total.Hippocampus \| BrainSegVol+CAT12+race+parent_educ | -0.0075 | 0.0045 | -1.6687 | 0.0969 | -0.0165 | 0.0014 | FALSE |
| Total.Hippocampus \| age+sex+TotalGrayVol+parent_educ | -0.0076 | 0.0043 | -1.7499 | 0.0818 | -0.0162 | 0.0010 | FALSE |
| Total.Hippocampus \| age+sex+TotalGrayVol+race | -0.0077 | 0.0044 | -1.7513 | 0.0816 | -0.0164 | 0.0010 | FALSE |
| Total.Hippocampus \| age+sex+BrainSegVol+CAT12+race+parent_educ | -0.0077 | 0.0044 | -1.7581 | 0.0805 | -0.0164 | 0.0010 | FALSE |
| Total.Hippocampus \| TotalGrayVol | -0.0078 | 0.0042 | -1.8565 | 0.0649 | -0.0161 | 0.0005 | FALSE |
| Left.Hippocampus \| age+sex+CAT12+race+parent_educ | -0.0078 | 0.0075 | -1.0466 | 0.2967 | -0.0225 | 0.0069 | FALSE |
| Total.Hippocampus \| TotalGrayVol+CAT12 | -0.0078 | 0.0042 | -1.8619 | 0.0641 | -0.0161 | 0.0005 | FALSE |
| Total.Hippocampus \| age+sex+TotalGrayVol+CAT12+parent_educ | -0.0079 | 0.0043 | -1.8061 | 0.0725 | -0.0164 | 0.0007 | FALSE |
| Total.Hippocampus \| age+sex+TotalGrayVol+CAT12+race | -0.0079 | 0.0044 | -1.7979 | 0.0739 | -0.0166 | 0.0008 | FALSE |
| Left.Hippocampus \| age+sex+race+parent_educ | -0.0079 | 0.0074 | -1.0638 | 0.2889 | -0.0226 | 0.0068 | FALSE |
| Total.Hippocampus \| BrainSegVol+parent_educ | -0.0080 | 0.0044 | -1.8239 | 0.0698 | -0.0166 | 0.0007 | FALSE |
| Total.Hippocampus \| age+sex+BrainSegVol+race | -0.0080 | 0.0043 | -1.8501 | 0.0660 | -0.0166 | 0.0005 | FALSE |
| Total.Hippocampus \| BrainSegVol+CAT12+parent_educ | -0.0081 | 0.0044 | -1.8423 | 0.0670 | -0.0168 | 0.0006 | FALSE |
| Total.Hippocampus \| age+sex+BrainSegVol+parent_educ | -0.0081 | 0.0043 | -1.8945 | 0.0597 | -0.0166 | 0.0003 | FALSE |
| Total.Hippocampus \| BrainSegVol+race | -0.0083 | 0.0045 | -1.8666 | 0.0636 | -0.0171 | 0.0005 | FALSE |
| Total.Hippocampus \| age+sex+BrainSegVol+CAT12+race | -0.0084 | 0.0044 | -1.9213 | 0.0563 | -0.0169 | 0.0002 | FALSE |
| Total.Hippocampus \| BrainSegVol+CAT12+race | -0.0084 | 0.0045 | -1.8687 | 0.0633 | -0.0172 | 0.0005 | FALSE |
| Left.Hippocampus \| age+sex+EstimatedTotalIntraCranialVol+race+parent_educ | -0.0087 | 0.0082 | -1.0669 | 0.2875 | -0.0248 | 0.0074 | FALSE |
| Left.Hippocampus \| TotalGrayVol+race+parent_educ | -0.0089 | 0.0089 | -0.9932 | 0.3220 | -0.0265 | 0.0088 | FALSE |
| Left.Hippocampus \| TotalGrayVol+CAT12+race+parent_educ | -0.0090 | 0.0090 | -1.0016 | 0.3179 | -0.0267 | 0.0087 | FALSE |
| Left.Hippocampus \| age+sex+EstimatedTotalIntraCranialVol+CAT12+race+parent_educ | -0.0090 | 0.0082 | -1.1027 | 0.2717 | -0.0251 | 0.0071 | FALSE |
| Left.Hippocampus \| age+sex+CAT12+race | -0.0093 | 0.0072 | -1.2939 | 0.1974 | -0.0236 | 0.0049 | FALSE |
| Left.Hippocampus \| TotalGrayVol+parent_educ | -0.0094 | 0.0087 | -1.0838 | 0.2799 | -0.0265 | 0.0077 | FALSE |
| Left.Hippocampus \| age+sex+race | -0.0095 | 0.0072 | -1.3150 | 0.1902 | -0.0237 | 0.0047 | FALSE |
| Left.Hippocampus \| TotalGrayVol+CAT12+parent_educ | -0.0096 | 0.0087 | -1.0990 | 0.2732 | -0.0268 | 0.0076 | FALSE |
| Right.Hippocampus \| age+sex+CAT12+race+parent_educ | -0.0097 | 0.0066 | -1.4634 | 0.1452 | -0.0228 | 0.0034 | FALSE |
| Left.Hippocampus \| age+sex+CAT12+parent_educ | -0.0097 | 0.0072 | -1.3506 | 0.1785 | -0.0240 | 0.0045 | FALSE |
| Left.Hippocampus \| age+sex+EstimatedTotalIntraCranialVol+parent_educ | -0.0098 | 0.0080 | -1.2283 | 0.2209 | -0.0255 | 0.0059 | FALSE |
| Left.Hippocampus \| age+sex+parent_educ | -0.0098 | 0.0072 | -1.3600 | 0.1755 | -0.0240 | 0.0044 | FALSE |
| Right.Hippocampus \| age+sex+race+parent_educ | -0.0099 | 0.0066 | -1.5015 | 0.1350 | -0.0230 | 0.0031 | FALSE |
| Left.Hippocampus \| EstimatedTotalIntraCranialVol+race+parent_educ | -0.0101 | 0.0083 | -1.2149 | 0.2260 | -0.0264 | 0.0062 | FALSE |
| Left.Hippocampus \| EstimatedTotalIntraCranialVol+CAT12+race+parent_educ | -0.0102 | 0.0083 | -1.2230 | 0.2229 | -0.0266 | 0.0062 | FALSE |
| Left.Hippocampus \| age+sex+EstimatedTotalIntraCranialVol+CAT12+parent_educ | -0.0102 | 0.0080 | -1.2746 | 0.2041 | -0.0260 | 0.0056 | FALSE |
| Left.Hippocampus \| age+sex+EstimatedTotalIntraCranialVol+race | -0.0102 | 0.0080 | -1.2865 | 0.1999 | -0.0259 | 0.0055 | FALSE |
| Right.Hippocampus \| age+sex+CAT12+race | -0.0104 | 0.0065 | -1.5952 | 0.1124 | -0.0233 | 0.0025 | FALSE |
| Left.Hippocampus \| age+sex+EstimatedTotalIntraCranialVol+CAT12+race | -0.0105 | 0.0080 | -1.3231 | 0.1875 | -0.0262 | 0.0052 | FALSE |
| Left.Hippocampus \| age+sex+TotalGrayVol+race+parent_educ | -0.0107 | 0.0088 | -1.2146 | 0.2261 | -0.0280 | 0.0067 | FALSE |
| Right.Hippocampus \| age+sex+race | -0.0107 | 0.0065 | -1.6415 | 0.1024 | -0.0236 | 0.0022 | FALSE |
| Left.Hippocampus \| EstimatedTotalIntraCranialVol+parent_educ | -0.0107 | 0.0081 | -1.3299 | 0.1852 | -0.0266 | 0.0052 | FALSE |
| Left.Hippocampus \| BrainSegVol+race+parent_educ | -0.0109 | 0.0090 | -1.2164 | 0.2254 | -0.0286 | 0.0068 | FALSE |
| Left.Hippocampus \| EstimatedTotalIntraCranialVol+CAT12+parent_educ | -0.0109 | 0.0081 | -1.3446 | 0.1804 | -0.0269 | 0.0051 | FALSE |
| Right.Hippocampus \| age+sex+EstimatedTotalIntraCranialVol+race+parent_educ | -0.0110 | 0.0072 | -1.5310 | 0.1276 | -0.0251 | 0.0032 | FALSE |
| Left.Hippocampus \| age+sex+TotalGrayVol+parent_educ | -0.0110 | 0.0086 | -1.2770 | 0.2032 | -0.0279 | 0.0060 | FALSE |
| Right.Hippocampus \| age+sex+EstimatedTotalIntraCranialVol+CAT12+race+parent_educ | -0.0110 | 0.0072 | -1.5390 | 0.1256 | -0.0251 | 0.0031 | FALSE |
| Left.Hippocampus \| BrainSegVol+CAT12+race+parent_educ | -0.0111 | 0.0090 | -1.2277 | 0.2212 | -0.0288 | 0.0067 | FALSE |
| Left.Hippocampus \| age+sex+TotalGrayVol+CAT12+race+parent_educ | -0.0112 | 0.0088 | -1.2696 | 0.2059 | -0.0285 | 0.0062 | FALSE |
| Left.Hippocampus \| age+sex+CAT12+race+parent_educ+puberty | -0.0112 | 0.0076 | -1.4822 | 0.1401 | -0.0261 | 0.0037 | FALSE |
| Left.Hippocampus \| TotalGrayVol+race | -0.0112 | 0.0088 | -1.2790 | 0.2025 | -0.0285 | 0.0061 | FALSE |
| Left.Hippocampus \| TotalGrayVol+CAT12+race | -0.0113 | 0.0088 | -1.2843 | 0.2007 | -0.0287 | 0.0061 | FALSE |
| Left.Hippocampus \| age+sex+race+parent_educ+puberty | -0.0114 | 0.0075 | -1.5044 | 0.1343 | -0.0263 | 0.0035 | FALSE |
| Left.Hippocampus \| age+sex+BrainSegVol+race+parent_educ | -0.0114 | 0.0087 | -1.3018 | 0.1947 | -0.0286 | 0.0059 | FALSE |
| Left.Hippocampus \| age+sex+TotalGrayVol+CAT12+parent_educ | -0.0115 | 0.0086 | -1.3405 | 0.1817 | -0.0285 | 0.0054 | FALSE |
| Right.Hippocampus \| age+sex+EstimatedTotalIntraCranialVol+race | -0.0116 | 0.0071 | -1.6324 | 0.1044 | -0.0256 | 0.0024 | FALSE |
| Left.Hippocampus \| BrainSegVol+parent_educ | -0.0116 | 0.0087 | -1.3308 | 0.1849 | -0.0288 | 0.0056 | FALSE |
| Right.Hippocampus \| age+sex+EstimatedTotalIntraCranialVol+CAT12+race | -0.0116 | 0.0071 | -1.6352 | 0.1038 | -0.0256 | 0.0024 | FALSE |
| Left.Hippocampus \| age+sex+EstimatedTotalIntraCranialVol+race+parent_educ+puberty | -0.0116 | 0.0082 | -1.4188 | 0.1578 | -0.0278 | 0.0045 | FALSE |
| Left.Hippocampus \| age+sex+EstimatedTotalIntraCranialVol+CAT12+race+parent_educ+puberty | -0.0118 | 0.0082 | -1.4420 | 0.1511 | -0.0280 | 0.0044 | FALSE |
| Left.Hippocampus \| BrainSegVol+CAT12+parent_educ | -0.0119 | 0.0088 | -1.3505 | 0.1785 | -0.0292 | 0.0055 | FALSE |
| Right.Hippocampus \| age+sex+CAT12+parent_educ | -0.0120 | 0.0064 | -1.8844 | 0.0611 | -0.0247 | 0.0006 | FALSE |
| Left.Hippocampus \| TotalGrayVol | -0.0121 | 0.0083 | -1.4666 | 0.1441 | -0.0284 | 0.0042 | FALSE |
| Left.Hippocampus \| age+sex+BrainSegVol+CAT12+race+parent_educ | -0.0121 | 0.0088 | -1.3841 | 0.1681 | -0.0294 | 0.0052 | FALSE |
| Left.Hippocampus \| age+sex+TotalGrayVol+race | -0.0121 | 0.0086 | -1.4148 | 0.1589 | -0.0291 | 0.0048 | FALSE |
| Left.Hippocampus \| age+sex+BrainSegVol+parent_educ | -0.0122 | 0.0086 | -1.4203 | 0.1572 | -0.0291 | 0.0047 | FALSE |
| Left.Hippocampus \| TotalGrayVol+CAT12 | -0.0122 | 0.0083 | -1.4748 | 0.1419 | -0.0286 | 0.0041 | FALSE |
| Right.Hippocampus \| age+sex+parent_educ | -0.0122 | 0.0064 | -1.9138 | 0.0572 | -0.0248 | 0.0004 | FALSE |
| Left.Hippocampus \| EstimatedTotalIntraCranialVol+race | -0.0124 | 0.0081 | -1.5387 | 0.1256 | -0.0284 | 0.0035 | FALSE |
| Left.Hippocampus \| age+sex+EstimatedTotalIntraCranialVol+parent_educ+puberty | -0.0125 | 0.0080 | -1.5643 | 0.1195 | -0.0282 | 0.0033 | FALSE |
| Left.Hippocampus \| age+sex+CAT12+race+puberty | -0.0125 | 0.0073 | -1.7151 | 0.0881 | -0.0269 | 0.0019 | FALSE |
| Left.Hippocampus \| EstimatedTotalIntraCranialVol+CAT12+race | -0.0125 | 0.0081 | -1.5424 | 0.1247 | -0.0286 | 0.0035 | FALSE |
| Right.Hippocampus \| TotalGrayVol+race+parent_educ | -0.0125 | 0.0079 | -1.5801 | 0.1159 | -0.0282 | 0.0031 | FALSE |
| Right.Hippocampus \| TotalGrayVol+CAT12+race+parent_educ | -0.0126 | 0.0080 | -1.5820 | 0.1154 | -0.0283 | 0.0031 | FALSE |
| Left.Hippocampus \| age+sex+race+puberty | -0.0127 | 0.0073 | -1.7396 | 0.0837 | -0.0271 | 0.0017 | FALSE |
| Left.Hippocampus \| age+sex+EstimatedTotalIntraCranialVol | -0.0127 | 0.0076 | -1.6745 | 0.0957 | -0.0276 | 0.0023 | FALSE |
| Left.Hippocampus \| age+sex+TotalGrayVol+CAT12+race | -0.0127 | 0.0086 | -1.4747 | 0.1421 | -0.0296 | 0.0043 | FALSE |
| Right.Hippocampus \| age+sex+EstimatedTotalIntraCranialVol+parent_educ | -0.0128 | 0.0070 | -1.8302 | 0.0689 | -0.0265 | 0.0010 | FALSE |
| Left.Hippocampus \| age+sex+EstimatedTotalIntraCranialVol+CAT12+parent_educ+puberty | -0.0128 | 0.0080 | -1.5980 | 0.1118 | -0.0286 | 0.0030 | FALSE |
| Left.Hippocampus \| age+sex+CAT12 | -0.0128 | 0.0067 | -1.8997 | 0.0590 | -0.0261 | 0.0005 | FALSE |
| Left.Hippocampus \| age+sex+CAT12+parent_educ+puberty | -0.0128 | 0.0073 | -1.7617 | 0.0798 | -0.0272 | 0.0015 | FALSE |
| Right.Hippocampus \| EstimatedTotalIntraCranialVol+race+parent_educ | -0.0128 | 0.0072 | -1.7859 | 0.0758 | -0.0270 | 0.0013 | FALSE |
| Left.Hippocampus \| age+sex+EstimatedTotalIntraCranialVol+CAT12 | -0.0129 | 0.0076 | -1.7018 | 0.0904 | -0.0278 | 0.0021 | FALSE |
| Left.Hippocampus \| age+sex+parent_educ+puberty | -0.0129 | 0.0073 | -1.7751 | 0.0775 | -0.0272 | 0.0014 | FALSE |
| Right.Hippocampus \| EstimatedTotalIntraCranialVol+CAT12+race+parent_educ | -0.0129 | 0.0072 | -1.7884 | 0.0754 | -0.0272 | 0.0013 | FALSE |
| Right.Hippocampus \| age+sex+EstimatedTotalIntraCranialVol+CAT12+parent_educ | -0.0129 | 0.0070 | -1.8510 | 0.0658 | -0.0267 | 0.0009 | FALSE |
| Left.Hippocampus \| age+sex+EstimatedTotalIntraCranialVol+race+puberty | -0.0129 | 0.0080 | -1.6211 | 0.1068 | -0.0287 | 0.0028 | FALSE |
| Right.Hippocampus \| TotalGrayVol+race | -0.0129 | 0.0079 | -1.6340 | 0.1040 | -0.0286 | 0.0027 | FALSE |
| Right.Hippocampus \| TotalGrayVol+CAT12+race | -0.0130 | 0.0079 | -1.6329 | 0.1042 | -0.0287 | 0.0027 | FALSE |
| Left.Hippocampus \| age+sex+BrainSegVol+race | -0.0130 | 0.0085 | -1.5204 | 0.1302 | -0.0298 | 0.0039 | FALSE |
| Left.Hippocampus \| age+sex+BrainSegVol+CAT12+parent_educ | -0.0130 | 0.0086 | -1.5116 | 0.1324 | -0.0300 | 0.0040 | FALSE |
| Left.Hippocampus \| age+sex | -0.0131 | 0.0067 | -1.9417 | 0.0536 | -0.0264 | 0.0002 | FALSE |
| Left.Hippocampus \| TotalGrayVol+race+parent_educ+puberty | -0.0131 | 0.0089 | -1.4710 | 0.1431 | -0.0307 | 0.0045 | FALSE |
| Left.Hippocampus \| age+sex+EstimatedTotalIntraCranialVol+CAT12+race+puberty | -0.0132 | 0.0080 | -1.6464 | 0.1015 | -0.0289 | 0.0026 | FALSE |
| Left.Hippocampus \| TotalGrayVol+CAT12+race+parent_educ+puberty | -0.0132 | 0.0090 | -1.4774 | 0.1414 | -0.0309 | 0.0044 | FALSE |
| Left.Hippocampus \| TotalGrayVol+parent_educ+puberty | -0.0134 | 0.0087 | -1.5456 | 0.1239 | -0.0304 | 0.0037 | FALSE |
| Left.Hippocampus \| BrainSegVol+race | -0.0134 | 0.0088 | -1.5321 | 0.1273 | -0.0307 | 0.0039 | FALSE |
| Right.Hippocampus \| TotalGrayVol+parent_educ | -0.0134 | 0.0077 | -1.7472 | 0.0823 | -0.0286 | 0.0017 | FALSE |
| Left.Hippocampus \| EstimatedTotalIntraCranialVol+race+parent_educ+puberty | -0.0135 | 0.0082 | -1.6386 | 0.1031 | -0.0297 | 0.0028 | FALSE |
| Left.Hippocampus \| age+sex+TotalGrayVol | -0.0135 | 0.0082 | -1.6547 | 0.0996 | -0.0296 | 0.0026 | FALSE |
| Right.Hippocampus \| TotalGrayVol+CAT12+parent_educ | -0.0135 | 0.0077 | -1.7539 | 0.0811 | -0.0287 | 0.0017 | FALSE |
| Left.Hippocampus \| TotalGrayVol+CAT12+parent_educ+puberty | -0.0135 | 0.0087 | -1.5584 | 0.1209 | -0.0307 | 0.0036 | FALSE |
| Left.Hippocampus \| BrainSegVol+CAT12+race | -0.0136 | 0.0088 | -1.5358 | 0.1263 | -0.0310 | 0.0039 | FALSE |
| Right.Hippocampus \| age+sex+TotalGrayVol+race+parent_educ | -0.0136 | 0.0078 | -1.7354 | 0.0844 | -0.0290 | 0.0019 | FALSE |
| Left.Hippocampus \| EstimatedTotalIntraCranialVol | -0.0136 | 0.0077 | -1.7727 | 0.0778 | -0.0287 | 0.0015 | FALSE |
| Left.Hippocampus \| EstimatedTotalIntraCranialVol+CAT12+race+parent_educ+puberty | -0.0136 | 0.0083 | -1.6447 | 0.1018 | -0.0299 | 0.0027 | FALSE |
| Right.Hippocampus \| EstimatedTotalIntraCranialVol+race | -0.0136 | 0.0072 | -1.9016 | 0.0588 | -0.0277 | 0.0005 | FALSE |
| Right.Hippocampus \| EstimatedTotalIntraCranialVol+CAT12+race | -0.0136 | 0.0072 | -1.8999 | 0.0591 | -0.0278 | 0.0005 | FALSE |
| Left.Hippocampus \| EstimatedTotalIntraCranialVol+CAT12 | -0.0137 | 0.0077 | -1.7802 | 0.0766 | -0.0288 | 0.0015 | FALSE |
| Left.Hippocampus \| age+sex+TotalGrayVol+race+parent_educ+puberty | -0.0137 | 0.0088 | -1.5545 | 0.1219 | -0.0311 | 0.0037 | FALSE |
| Left.Hippocampus \| EstimatedTotalIntraCranialVol+parent_educ+puberty | -0.0137 | 0.0080 | -1.7193 | 0.0873 | -0.0295 | 0.0020 | FALSE |
| Left.Hippocampus \| age+sex+BrainSegVol+CAT12+race | -0.0138 | 0.0086 | -1.6061 | 0.1100 | -0.0307 | 0.0031 | FALSE |
| Right.Hippocampus \| age+sex+TotalGrayVol+CAT12+race+parent_educ | -0.0138 | 0.0078 | -1.7643 | 0.0794 | -0.0292 | 0.0016 | FALSE |
| Right.Hippocampus \| age+sex+BrainSegVol+race+parent_educ | -0.0138 | 0.0077 | -1.7993 | 0.0737 | -0.0290 | 0.0013 | FALSE |
| Left.Hippocampus \| EstimatedTotalIntraCranialVol+CAT12+parent_educ+puberty | -0.0139 | 0.0080 | -1.7315 | 0.0851 | -0.0298 | 0.0019 | FALSE |
| Left.Hippocampus \| age+sex+TotalGrayVol+CAT12 | -0.0139 | 0.0082 | -1.7029 | 0.0902 | -0.0301 | 0.0022 | FALSE |
| Left.Hippocampus \| age+sex+TotalGrayVol+parent_educ+puberty | -0.0140 | 0.0086 | -1.6293 | 0.1050 | -0.0310 | 0.0030 | FALSE |
| Right.Hippocampus \| age+sex+TotalGrayVol+race | -0.0141 | 0.0078 | -1.8069 | 0.0725 | -0.0294 | 0.0013 | FALSE |
| Left.Hippocampus \| age+sex+TotalGrayVol+CAT12+race+parent_educ+puberty | -0.0141 | 0.0088 | -1.5949 | 0.1126 | -0.0315 | 0.0033 | FALSE |
| Right.Hippocampus \| age+sex+EstimatedTotalIntraCranialVol+CAT12+race+parent_educ+puberty | -0.0142 | 0.0072 | -1.9609 | 0.0515 | -0.0284 | 0.0001 | FALSE |
| Right.Hippocampus \| age+sex+EstimatedTotalIntraCranialVol+race+parent_educ+puberty | -0.0142 | 0.0072 | -1.9650 | 0.0510 | -0.0284 | 0.0001 | FALSE |
| Right.Hippocampus \| age+sex+BrainSegVol+CAT12+race+parent_educ | -0.0142 | 0.0077 | -1.8505 | 0.0659 | -0.0294 | 0.0009 | FALSE |
| Right.Hippocampus \| age+sex+TotalGrayVol+CAT12+race | -0.0143 | 0.0078 | -1.8341 | 0.0683 | -0.0296 | 0.0011 | FALSE |
| Right.Hippocampus \| BrainSegVol+race+parent_educ | -0.0143 | 0.0078 | -1.8254 | 0.0696 | -0.0298 | 0.0012 | FALSE |
| Right.Hippocampus \| BrainSegVol+CAT12+race+parent_educ | -0.0144 | 0.0079 | -1.8318 | 0.0687 | -0.0300 | 0.0011 | FALSE |
| Right.Hippocampus \| age+sex+BrainSegVol+race | -0.0144 | 0.0076 | -1.8887 | 0.0606 | -0.0295 | 0.0006 | FALSE |
| Left.Hippocampus \| age+sex+BrainSegVol+race+parent_educ+puberty | -0.0145 | 0.0088 | -1.6474 | 0.1013 | -0.0318 | 0.0029 | FALSE |
| Left.Hippocampus \| age+sex+TotalGrayVol+CAT12+parent_educ+puberty | -0.0145 | 0.0086 | -1.6782 | 0.0950 | -0.0315 | 0.0025 | FALSE |
| Right.Hippocampus \| TotalGrayVol | -0.0145 | 0.0074 | -1.9584 | 0.0516 | -0.0291 | 0.0001 | FALSE |
| Left.Hippocampus \| BrainSegVol | -0.0146 | 0.0083 | -1.7517 | 0.0814 | -0.0309 | 0.0018 | FALSE |
| Right.Hippocampus \| TotalGrayVol+CAT12 | -0.0146 | 0.0074 | -1.9604 | 0.0514 | -0.0292 | 0.0001 | FALSE |
| Left.Hippocampus \| BrainSegVol+CAT12 | -0.0147 | 0.0083 | -1.7653 | 0.0791 | -0.0312 | 0.0017 | FALSE |
| Right.Hippocampus \| age+sex+TotalGrayVol+parent_educ | -0.0148 | 0.0076 | -1.9379 | 0.0542 | -0.0298 | 0.0003 | FALSE |
| Right.Hippocampus \| age+sex+BrainSegVol+CAT12+race | -0.0148 | 0.0077 | -1.9353 | 0.0545 | -0.0299 | 0.0003 | FALSE |
| Right.Hippocampus \| BrainSegVol+race | -0.0150 | 0.0078 | -1.9054 | 0.0583 | -0.0303 | 0.0005 | FALSE |
| Left.Hippocampus \| age+sex+TotalGrayVol+race+puberty | -0.0149 | 0.0086 | -1.7374 | 0.0841 | -0.0319 | 0.0020 | FALSE |
| Left.Hippocampus \| BrainSegVol+race+parent_educ+puberty | -0.0150 | 0.0089 | -1.6793 | 0.0949 | -0.0325 | 0.0026 | FALSE |
| Right.Hippocampus \| BrainSegVol+CAT12+race | -0.0150 | 0.0079 | -1.9044 | 0.0585 | -0.0305 | 0.0005 | FALSE |
| Left.Hippocampus \| TotalGrayVol+race+puberty | -0.0151 | 0.0087 | -1.7290 | 0.0855 | -0.0323 | 0.0021 | FALSE |
| Left.Hippocampus \| age+sex+BrainSegVol+CAT12+race+parent_educ+puberty | -0.0151 | 0.0088 | -1.7108 | 0.0889 | -0.0324 | 0.0023 | FALSE |
| Left.Hippocampus \| age+sex+BrainSegVol | -0.0151 | 0.0082 | -1.8475 | 0.0662 | -0.0312 | 0.0010 | FALSE |
| Left.Hippocampus \| BrainSegVol+CAT12+race+parent_educ+puberty | -0.0151 | 0.0090 | -1.6884 | 0.0931 | -0.0328 | 0.0026 | FALSE |
| Left.Hippocampus \| TotalGrayVol+CAT12+race+puberty | -0.0152 | 0.0088 | -1.7340 | 0.0847 | -0.0324 | 0.0021 | FALSE |
| Left.Hippocampus \| age+sex+BrainSegVol+parent_educ+puberty | -0.0152 | 0.0086 | -1.7767 | 0.0773 | -0.0322 | 0.0017 | FALSE |
| Left.Hippocampus \| age+sex+TotalGrayVol+CAT12+race+puberty | -0.0154 | 0.0086 | -1.7833 | 0.0763 | -0.0324 | 0.0016 | FALSE |
| Left.Hippocampus \| EstimatedTotalIntraCranialVol+race+puberty | -0.0154 | 0.0080 | -1.9240 | 0.0559 | -0.0312 | 0.0004 | FALSE |
| Left.Hippocampus \| BrainSegVol+parent_educ+puberty | -0.0154 | 0.0087 | -1.7808 | 0.0766 | -0.0326 | 0.0017 | FALSE |
| Left.Hippocampus \| EstimatedTotalIntraCranialVol+CAT12+race+puberty | -0.0155 | 0.0080 | -1.9276 | 0.0555 | -0.0314 | 0.0004 | FALSE |
| Left.Hippocampus \| BrainSegVol+CAT12+parent_educ+puberty | -0.0157 | 0.0087 | -1.7978 | 0.0739 | -0.0329 | 0.0015 | FALSE |
| Left.Hippocampus \| age+sex+BrainSegVol+CAT12 | -0.0158 | 0.0082 | -1.9258 | 0.0556 | -0.0319 | 0.0004 | FALSE |
| Left.Hippocampus \| age+sex+BrainSegVol+race+puberty | -0.0158 | 0.0086 | -1.8502 | 0.0660 | -0.0327 | 0.0011 | FALSE |
| Left.Hippocampus \| age+sex+BrainSegVol+CAT12+parent_educ+puberty | -0.0159 | 0.0086 | -1.8494 | 0.0660 | -0.0330 | 0.0011 | FALSE |
| Left.Hippocampus \| TotalGrayVol+puberty | -0.0160 | 0.0082 | -1.9438 | 0.0534 | -0.0322 | 0.0002 | FALSE |
| Left.Hippocampus \| TotalGrayVol+CAT12+puberty | -0.0161 | 0.0082 | -1.9557 | 0.0520 | -0.0324 | 0.0001 | FALSE |
| Left.Hippocampus \| age+sex+BrainSegVol+CAT12+race+puberty | -0.0165 | 0.0086 | -1.9181 | 0.0567 | -0.0334 | 0.0005 | FALSE |
| Left.Hippocampus \| BrainSegVol+race+puberty | -0.0171 | 0.0087 | -1.9669 | 0.0507 | -0.0342 | 0.0001 | FALSE |
| Left.Hippocampus \| BrainSegVol+CAT12+race+puberty | -0.0172 | 0.0087 | -1.9713 | 0.0503 | -0.0345 | 0.0000 | FALSE |
| Total.Hippocampus \| age+sex+CAT12+race+puberty | -0.0075 | 0.0037 | -2.0310 | 0.0438 | -0.0147 | -0.0002 | TRUE |
| Total.Hippocampus \| age+sex+CAT12 | -0.0075 | 0.0034 | -2.2476 | 0.0257 | -0.0141 | -0.0009 | TRUE |
| Total.Hippocampus \| age+sex+race+puberty | -0.0076 | 0.0037 | -2.0713 | 0.0398 | -0.0148 | -0.0004 | TRUE |
| Total.Hippocampus \| age+sex | -0.0077 | 0.0033 | -2.2969 | 0.0227 | -0.0143 | -0.0011 | TRUE |
| Total.Hippocampus \| age+sex+EstimatedTotalIntraCranialVol | -0.0078 | 0.0038 | -2.0704 | 0.0398 | -0.0153 | -0.0004 | TRUE |
| Total.Hippocampus \| age+sex+EstimatedTotalIntraCranialVol+CAT12 | -0.0079 | 0.0038 | -2.0910 | 0.0379 | -0.0154 | -0.0004 | TRUE |
| Total.Hippocampus \| age+sex+CAT12+parent_educ+puberty | -0.0079 | 0.0036 | -2.2048 | 0.0287 | -0.0150 | -0.0008 | TRUE |
| Total.Hippocampus \| age+sex+EstimatedTotalIntraCranialVol+race+puberty | -0.0080 | 0.0040 | -1.9829 | 0.0489 | -0.0159 | 0.0000 | TRUE |
| Total.Hippocampus \| age+sex+EstimatedTotalIntraCranialVol+CAT12+race+puberty | -0.0080 | 0.0040 | -1.9911 | 0.0480 | -0.0160 | -0.0001 | TRUE |
| Total.Hippocampus \| age+sex+parent_educ+puberty | -0.0080 | 0.0036 | -2.2308 | 0.0269 | -0.0151 | -0.0009 | TRUE |
| Total.Hippocampus \| age+sex+EstimatedTotalIntraCranialVol+parent_educ+puberty | -0.0082 | 0.0040 | -2.0608 | 0.0408 | -0.0160 | -0.0003 | TRUE |
| Total.Hippocampus \| EstimatedTotalIntraCranialVol | -0.0082 | 0.0038 | -2.1619 | 0.0319 | -0.0157 | -0.0007 | TRUE |
| Total.Hippocampus \| age+sex+EstimatedTotalIntraCranialVol+CAT12+parent_educ+puberty | -0.0083 | 0.0040 | -2.0825 | 0.0387 | -0.0161 | -0.0004 | TRUE |
| Total.Hippocampus \| EstimatedTotalIntraCranialVol+CAT12 | -0.0083 | 0.0038 | -2.1666 | 0.0315 | -0.0158 | -0.0007 | TRUE |
| Total.Hippocampus \| age+sex+BrainSegVol+CAT12+parent_educ | -0.0085 | 0.0043 | -1.9785 | 0.0494 | -0.0170 | 0.0000 | TRUE |
| Total.Hippocampus \| EstimatedTotalIntraCranialVol+race+parent_educ+puberty | -0.0086 | 0.0041 | -2.1154 | 0.0358 | -0.0166 | -0.0006 | TRUE |
| Total.Hippocampus \| EstimatedTotalIntraCranialVol+CAT12+race+parent_educ+puberty | -0.0087 | 0.0041 | -2.1188 | 0.0355 | -0.0167 | -0.0006 | TRUE |
| Total.Hippocampus \| age+sex+TotalGrayVol | -0.0087 | 0.0042 | -2.0973 | 0.0373 | -0.0169 | -0.0005 | TRUE |
| Total.Hippocampus \| race+parent_educ | -0.0088 | 0.0034 | -2.5842 | 0.0106 | -0.0156 | -0.0021 | TRUE |
| Total.Hippocampus \| CAT12+race+parent_educ | -0.0088 | 0.0034 | -2.5842 | 0.0106 | -0.0156 | -0.0021 | TRUE |
| Total.Hippocampus \| EstimatedTotalIntraCranialVol+parent_educ+puberty | -0.0089 | 0.0040 | -2.2533 | 0.0254 | -0.0167 | -0.0011 | TRUE |
| Total.Hippocampus \| age+sex+TotalGrayVol+CAT12 | -0.0089 | 0.0042 | -2.1436 | 0.0333 | -0.0171 | -0.0007 | TRUE |
| Total.Hippocampus \| TotalGrayVol+race+parent_educ+puberty | -0.0090 | 0.0045 | -1.9786 | 0.0494 | -0.0179 | 0.0000 | TRUE |
| Total.Hippocampus \| EstimatedTotalIntraCranialVol+CAT12+parent_educ+puberty | -0.0090 | 0.0040 | -2.2622 | 0.0249 | -0.0168 | -0.0011 | TRUE |
| Total.Hippocampus \| TotalGrayVol+CAT12+race+parent_educ+puberty | -0.0090 | 0.0045 | -1.9816 | 0.0491 | -0.0180 | 0.0000 | TRUE |
| Total.Hippocampus \| age+sex+TotalGrayVol+race+parent_educ+puberty | -0.0091 | 0.0045 | -2.0263 | 0.0443 | -0.0179 | -0.0002 | TRUE |
| Total.Hippocampus \| BrainSegVol | -0.0091 | 0.0042 | -2.1749 | 0.0308 | -0.0174 | -0.0008 | TRUE |
| Total.Hippocampus \| BrainSegVol+CAT12 | -0.0092 | 0.0042 | -2.1872 | 0.0299 | -0.0175 | -0.0009 | TRUE |
| Total.Hippocampus \| age+sex+TotalGrayVol+CAT12+race+parent_educ+puberty | -0.0092 | 0.0045 | -2.0551 | 0.0414 | -0.0181 | -0.0004 | TRUE |
| Total.Hippocampus \| TotalGrayVol+parent_educ+puberty | -0.0093 | 0.0044 | -2.1128 | 0.0360 | -0.0179 | -0.0006 | TRUE |
| Total.Hippocampus \| EstimatedTotalIntraCranialVol+race+puberty | -0.0093 | 0.0040 | -2.3268 | 0.0211 | -0.0172 | -0.0014 | TRUE |
| Total.Hippocampus \| age+sex+CAT12+puberty | -0.0093 | 0.0034 | -2.7512 | 0.0065 | -0.0160 | -0.0026 | TRUE |
| Total.Hippocampus \| TotalGrayVol+CAT12+parent_educ+puberty | -0.0093 | 0.0044 | -2.1216 | 0.0352 | -0.0180 | -0.0007 | TRUE |
| Total.Hippocampus \| EstimatedTotalIntraCranialVol+CAT12+race+puberty | -0.0094 | 0.0040 | -2.3272 | 0.0211 | -0.0173 | -0.0014 | TRUE |
| Total.Hippocampus \| age+sex+BrainSegVol+race+parent_educ+puberty | -0.0094 | 0.0044 | -2.1127 | 0.0361 | -0.0181 | -0.0006 | TRUE |
| Total.Hippocampus \| parent_educ | -0.0094 | 0.0033 | -2.8684 | 0.0046 | -0.0159 | -0.0029 | TRUE |
| Total.Hippocampus \| age+sex+EstimatedTotalIntraCranialVol+puberty | -0.0094 | 0.0038 | -2.4972 | 0.0134 | -0.0169 | -0.0020 | TRUE |
| Total.Hippocampus \| age+sex+BrainSegVol | -0.0095 | 0.0041 | -2.2959 | 0.0228 | -0.0176 | -0.0013 | TRUE |
| Total.Hippocampus \| age+sex+puberty | -0.0095 | 0.0034 | -2.8020 | 0.0056 | -0.0161 | -0.0028 | TRUE |
| Total.Hippocampus \| CAT12+parent_educ | -0.0095 | 0.0033 | -2.8723 | 0.0046 | -0.0160 | -0.0030 | TRUE |
| Total.Hippocampus \| age+sex+EstimatedTotalIntraCranialVol+CAT12+puberty | -0.0095 | 0.0038 | -2.5110 | 0.0129 | -0.0170 | -0.0020 | TRUE |
| Total.Hippocampus \| age+sex+TotalGrayVol+parent_educ+puberty | -0.0096 | 0.0044 | -2.1920 | 0.0297 | -0.0182 | -0.0010 | TRUE |
| Total.Hippocampus \| age+sex+TotalGrayVol+race+puberty | -0.0096 | 0.0044 | -2.1630 | 0.0319 | -0.0183 | -0.0008 | TRUE |
| Total.Hippocampus \| age+sex+BrainSegVol+CAT12+race+parent_educ+puberty | -0.0096 | 0.0044 | -2.1641 | 0.0318 | -0.0184 | -0.0008 | TRUE |
| Total.Hippocampus \| TotalGrayVol+race+puberty | -0.0096 | 0.0045 | -2.1541 | 0.0326 | -0.0185 | -0.0008 | TRUE |
| Total.Hippocampus \| TotalGrayVol+CAT12+race+puberty | -0.0097 | 0.0045 | -2.1553 | 0.0325 | -0.0186 | -0.0008 | TRUE |
| Total.Hippocampus \| age+sex+TotalGrayVol+CAT12+race+puberty | -0.0097 | 0.0044 | -2.1944 | 0.0295 | -0.0185 | -0.0010 | TRUE |
| Total.Hippocampus \| age+sex+TotalGrayVol+CAT12+parent_educ+puberty | -0.0097 | 0.0044 | -2.2319 | 0.0269 | -0.0184 | -0.0011 | TRUE |
| Total.Hippocampus \| age+sex+BrainSegVol+CAT12 | -0.0098 | 0.0041 | -2.3722 | 0.0187 | -0.0179 | -0.0017 | TRUE |
| Total.Hippocampus \| age+sex+BrainSegVol+race+puberty | -0.0099 | 0.0044 | -2.2705 | 0.0244 | -0.0185 | -0.0013 | TRUE |
| Total.Hippocampus \| BrainSegVol+race+parent_educ+puberty | -0.0099 | 0.0045 | -2.2138 | 0.0281 | -0.0188 | -0.0011 | TRUE |
| Total.Hippocampus \| BrainSegVol+CAT12+race+parent_educ+puberty | -0.0100 | 0.0045 | -2.2214 | 0.0276 | -0.0189 | -0.0011 | TRUE |
| Total.Hippocampus \| race | -0.0100 | 0.0033 | -3.0394 | 0.0027 | -0.0165 | -0.0035 | TRUE |
| Total.Hippocampus \| CAT12+race | -0.0100 | 0.0033 | -3.0368 | 0.0027 | -0.0166 | -0.0035 | TRUE |
| Total.Hippocampus \| EstimatedTotalIntraCranialVol+puberty | -0.0100 | 0.0038 | -2.6629 | 0.0084 | -0.0175 | -0.0026 | TRUE |
| Total.Hippocampus \| EstimatedTotalIntraCranialVol+CAT12+puberty | -0.0101 | 0.0038 | -2.6708 | 0.0082 | -0.0176 | -0.0026 | TRUE |
| Total.Hippocampus \| age+sex+BrainSegVol+parent_educ+puberty | -0.0101 | 0.0043 | -2.3415 | 0.0203 | -0.0186 | -0.0016 | TRUE |
| Total.Hippocampus \| age+sex+BrainSegVol+CAT12+race+puberty | -0.0102 | 0.0044 | -2.3225 | 0.0214 | -0.0188 | -0.0015 | TRUE |
| Total.Hippocampus \| TotalGrayVol+puberty | -0.0103 | 0.0042 | -2.4622 | 0.0147 | -0.0186 | -0.0021 | TRUE |
| Total.Hippocampus \| BrainSegVol+parent_educ+puberty | -0.0104 | 0.0044 | -2.3794 | 0.0184 | -0.0190 | -0.0018 | TRUE |
| Total.Hippocampus \| TotalGrayVol+CAT12+puberty | -0.0104 | 0.0042 | -2.4710 | 0.0144 | -0.0187 | -0.0021 | TRUE |
| Total.Hippocampus \| age+sex+BrainSegVol+CAT12+parent_educ+puberty | -0.0104 | 0.0043 | -2.4054 | 0.0172 | -0.0190 | -0.0019 | TRUE |
| Total.Hippocampus \| race+parent_educ+puberty | -0.0104 | 0.0034 | -3.0790 | 0.0024 | -0.0171 | -0.0037 | TRUE |
| Total.Hippocampus \| CAT12+race+parent_educ+puberty | -0.0105 | 0.0034 | -3.0771 | 0.0024 | -0.0172 | -0.0037 | TRUE |
| Total.Hippocampus \| BrainSegVol+CAT12+parent_educ+puberty | -0.0105 | 0.0044 | -2.3949 | 0.0176 | -0.0191 | -0.0018 | TRUE |
| Total.Hippocampus \| age+sex+TotalGrayVol+puberty | -0.0106 | 0.0042 | -2.5497 | 0.0116 | -0.0189 | -0.0024 | TRUE |
| Total.Hippocampus \| BrainSegVol+race+puberty | -0.0107 | 0.0044 | -2.4187 | 0.0166 | -0.0194 | -0.0020 | TRUE |
| Total.Hippocampus \| BrainSegVol+CAT12+race+puberty | -0.0108 | 0.0045 | -2.4209 | 0.0165 | -0.0196 | -0.0020 | TRUE |
| Total.Hippocampus \| age+sex+TotalGrayVol+CAT12+puberty | -0.0108 | 0.0042 | -2.5881 | 0.0104 | -0.0190 | -0.0026 | TRUE |
| Total.Hippocampus \| parent_educ+puberty | -0.0108 | 0.0033 | -3.3231 | 0.0011 | -0.0172 | -0.0044 | TRUE |
| Total.Hippocampus \| CAT12+parent_educ+puberty | -0.0109 | 0.0033 | -3.3251 | 0.0011 | -0.0173 | -0.0044 | TRUE |
| Total.Hippocampus \| CAT12 | -0.0110 | 0.0031 | -3.5709 | 0.0004 | -0.0170 | -0.0049 | TRUE |
| Total.Hippocampus \| unadjusted | -0.0110 | 0.0031 | -3.5791 | 0.0004 | -0.0170 | -0.0049 | TRUE |
| Total.Hippocampus \| race+puberty | -0.0113 | 0.0033 | -3.4827 | 0.0006 | -0.0178 | -0.0049 | TRUE |
| Total.Hippocampus \| CAT12+race+puberty | -0.0114 | 0.0033 | -3.4793 | 0.0006 | -0.0178 | -0.0049 | TRUE |
| Total.Hippocampus \| age+sex+BrainSegVol+puberty | -0.0114 | 0.0041 | -2.7556 | 0.0064 | -0.0195 | -0.0032 | TRUE |
| Total.Hippocampus \| BrainSegVol+puberty | -0.0115 | 0.0042 | -2.7699 | 0.0062 | -0.0198 | -0.0033 | TRUE |
| Total.Hippocampus \| BrainSegVol+CAT12+puberty | -0.0117 | 0.0042 | -2.7893 | 0.0058 | -0.0199 | -0.0034 | TRUE |
| Total.Hippocampus \| age+sex+BrainSegVol+CAT12+puberty | -0.0117 | 0.0041 | -2.8221 | 0.0053 | -0.0199 | -0.0035 | TRUE |
| Total.Hippocampus \| puberty | -0.0122 | 0.0030 | -4.0233 | 0.0001 | -0.0181 | -0.0062 | TRUE |
| Total.Hippocampus \| CAT12+puberty | -0.0122 | 0.0030 | -4.0167 | 0.0001 | -0.0181 | -0.0062 | TRUE |
| Right.Hippocampus \| age+sex+CAT12+race+parent_educ+puberty | -0.0133 | 0.0067 | -1.9756 | 0.0498 | -0.0266 | 0.0000 | TRUE |
| Right.Hippocampus \| age+sex+race+parent_educ+puberty | -0.0136 | 0.0067 | -2.0191 | 0.0450 | -0.0268 | -0.0003 | TRUE |
| Right.Hippocampus \| EstimatedTotalIntraCranialVol+parent_educ | -0.0138 | 0.0070 | -1.9751 | 0.0497 | -0.0277 | 0.0000 | TRUE |
| Right.Hippocampus \| EstimatedTotalIntraCranialVol+CAT12+parent_educ | -0.0139 | 0.0070 | -1.9821 | 0.0490 | -0.0278 | -0.0001 | TRUE |
| Right.Hippocampus \| age+sex+CAT12+race+puberty | -0.0140 | 0.0066 | -2.1068 | 0.0366 | -0.0271 | -0.0009 | TRUE |
| Right.Hippocampus \| age+sex+race+puberty | -0.0143 | 0.0066 | -2.1569 | 0.0324 | -0.0273 | -0.0012 | TRUE |
| Right.Hippocampus \| age+sex+CAT12 | -0.0143 | 0.0061 | -2.3554 | 0.0195 | -0.0263 | -0.0023 | TRUE |
| Right.Hippocampus \| age+sex | -0.0146 | 0.0061 | -2.4070 | 0.0170 | -0.0266 | -0.0026 | TRUE |
| Right.Hippocampus \| age+sex+EstimatedTotalIntraCranialVol | -0.0147 | 0.0067 | -2.1896 | 0.0298 | -0.0279 | -0.0015 | TRUE |
| Right.Hippocampus \| age+sex+EstimatedTotalIntraCranialVol+CAT12+race+puberty | -0.0147 | 0.0072 | -2.0582 | 0.0410 | -0.0288 | -0.0006 | TRUE |
| Right.Hippocampus \| age+sex+EstimatedTotalIntraCranialVol+race+puberty | -0.0148 | 0.0071 | -2.0661 | 0.0403 | -0.0289 | -0.0007 | TRUE |
| Right.Hippocampus \| age+sex+EstimatedTotalIntraCranialVol+CAT12 | -0.0148 | 0.0067 | -2.2016 | 0.0289 | -0.0280 | -0.0015 | TRUE |
| Right.Hippocampus \| age+sex+TotalGrayVol+CAT12+parent_educ | -0.0151 | 0.0076 | -1.9786 | 0.0494 | -0.0302 | 0.0000 | TRUE |
| Left.Hippocampus \| age+sex+EstimatedTotalIntraCranialVol+puberty | -0.0153 | 0.0076 | -2.0155 | 0.0453 | -0.0302 | -0.0003 | TRUE |
| Right.Hippocampus \| EstimatedTotalIntraCranialVol | -0.0153 | 0.0068 | -2.2640 | 0.0247 | -0.0286 | -0.0020 | TRUE |
| Right.Hippocampus \| EstimatedTotalIntraCranialVol+CAT12 | -0.0153 | 0.0068 | -2.2654 | 0.0246 | -0.0287 | -0.0020 | TRUE |
| Right.Hippocampus \| BrainSegVol+parent_educ | -0.0154 | 0.0076 | -2.0181 | 0.0450 | -0.0304 | -0.0003 | TRUE |
| Right.Hippocampus \| age+sex+CAT12+parent_educ+puberty | -0.0154 | 0.0065 | -2.3851 | 0.0181 | -0.0281 | -0.0027 | TRUE |
| Left.Hippocampus \| age+sex+EstimatedTotalIntraCranialVol+CAT12+puberty | -0.0154 | 0.0076 | -2.0372 | 0.0430 | -0.0304 | -0.0005 | TRUE |
| Right.Hippocampus \| age+sex+BrainSegVol+parent_educ | -0.0155 | 0.0075 | -2.0638 | 0.0404 | -0.0303 | -0.0007 | TRUE |
| Left.Hippocampus \| race+parent_educ | -0.0155 | 0.0070 | -2.2049 | 0.0287 | -0.0295 | -0.0016 | TRUE |
| Right.Hippocampus \| BrainSegVol+CAT12+parent_educ | -0.0156 | 0.0077 | -2.0310 | 0.0437 | -0.0307 | -0.0004 | TRUE |
| Right.Hippocampus \| age+sex+parent_educ+puberty | -0.0156 | 0.0064 | -2.4195 | 0.0165 | -0.0283 | -0.0029 | TRUE |
| Left.Hippocampus \| CAT12+race+parent_educ | -0.0156 | 0.0071 | -2.2081 | 0.0285 | -0.0296 | -0.0017 | TRUE |
| Left.Hippocampus \| age+sex+CAT12+puberty | -0.0158 | 0.0068 | -2.3220 | 0.0213 | -0.0292 | -0.0024 | TRUE |
| Right.Hippocampus \| age+sex+EstimatedTotalIntraCranialVol+parent_educ+puberty | -0.0158 | 0.0070 | -2.2618 | 0.0249 | -0.0296 | -0.0020 | TRUE |
| Right.Hippocampus \| age+sex+EstimatedTotalIntraCranialVol+CAT12+parent_educ+puberty | -0.0159 | 0.0070 | -2.2700 | 0.0244 | -0.0297 | -0.0021 | TRUE |
| Right.Hippocampus \| age+sex+BrainSegVol+CAT12+parent_educ | -0.0160 | 0.0075 | -2.1276 | 0.0347 | -0.0309 | -0.0012 | TRUE |
| Left.Hippocampus \| age+sex+puberty | -0.0160 | 0.0068 | -2.3643 | 0.0191 | -0.0294 | -0.0027 | TRUE |
| Right.Hippocampus \| age+sex+TotalGrayVol | -0.0163 | 0.0074 | -2.2134 | 0.0280 | -0.0308 | -0.0018 | TRUE |
| Right.Hippocampus \| EstimatedTotalIntraCranialVol+race+parent_educ+puberty | -0.0163 | 0.0071 | -2.2858 | 0.0235 | -0.0304 | -0.0022 | TRUE |
| Right.Hippocampus \| EstimatedTotalIntraCranialVol+CAT12+race+parent_educ+puberty | -0.0164 | 0.0072 | -2.2857 | 0.0235 | -0.0305 | -0.0022 | TRUE |
| Left.Hippocampus \| age+sex+TotalGrayVol+puberty | -0.0164 | 0.0082 | -2.0098 | 0.0459 | -0.0325 | -0.0003 | TRUE |
| Left.Hippocampus \| EstimatedTotalIntraCranialVol+puberty | -0.0164 | 0.0076 | -2.1699 | 0.0312 | -0.0314 | -0.0015 | TRUE |
| Right.Hippocampus \| race+parent_educ | -0.0165 | 0.0061 | -2.7016 | 0.0076 | -0.0285 | -0.0044 | TRUE |
| Right.Hippocampus \| CAT12+race+parent_educ | -0.0165 | 0.0061 | -2.6988 | 0.0076 | -0.0286 | -0.0044 | TRUE |
| Right.Hippocampus \| age+sex+TotalGrayVol+CAT12 | -0.0166 | 0.0074 | -2.2506 | 0.0255 | -0.0311 | -0.0020 | TRUE |
| Left.Hippocampus \| EstimatedTotalIntraCranialVol+CAT12+puberty | -0.0166 | 0.0076 | -2.1811 | 0.0304 | -0.0316 | -0.0016 | TRUE |
| Right.Hippocampus \| BrainSegVol | -0.0166 | 0.0074 | -2.2619 | 0.0248 | -0.0311 | -0.0021 | TRUE |
| Left.Hippocampus \| parent_educ | -0.0167 | 0.0068 | -2.4508 | 0.0152 | -0.0301 | -0.0032 | TRUE |
| Left.Hippocampus \| CAT12+parent_educ | -0.0168 | 0.0068 | -2.4588 | 0.0149 | -0.0302 | -0.0033 | TRUE |
| Right.Hippocampus \| BrainSegVol+CAT12 | -0.0168 | 0.0074 | -2.2700 | 0.0243 | -0.0313 | -0.0022 | TRUE |
| Left.Hippocampus \| age+sex+TotalGrayVol+CAT12+puberty | -0.0168 | 0.0082 | -2.0513 | 0.0416 | -0.0329 | -0.0006 | TRUE |
| Right.Hippocampus \| age+sex+TotalGrayVol+race+parent_educ+puberty | -0.0170 | 0.0079 | -2.1606 | 0.0321 | -0.0325 | -0.0015 | TRUE |
| Right.Hippocampus \| TotalGrayVol+race+parent_educ+puberty | -0.0171 | 0.0079 | -2.1521 | 0.0328 | -0.0327 | -0.0014 | TRUE |
| Right.Hippocampus \| EstimatedTotalIntraCranialVol+race+puberty | -0.0171 | 0.0071 | -2.4062 | 0.0171 | -0.0311 | -0.0031 | TRUE |
| Right.Hippocampus \| EstimatedTotalIntraCranialVol+CAT12+race+puberty | -0.0171 | 0.0071 | -2.4032 | 0.0173 | -0.0311 | -0.0031 | TRUE |
| Right.Hippocampus \| TotalGrayVol+CAT12+race+parent_educ+puberty | -0.0171 | 0.0080 | -2.1513 | 0.0328 | -0.0328 | -0.0014 | TRUE |
| Right.Hippocampus \| EstimatedTotalIntraCranialVol+parent_educ+puberty | -0.0171 | 0.0069 | -2.4645 | 0.0146 | -0.0308 | -0.0034 | TRUE |
| Right.Hippocampus \| age+sex+TotalGrayVol+CAT12+race+parent_educ+puberty | -0.0171 | 0.0079 | -2.1742 | 0.0311 | -0.0327 | -0.0016 | TRUE |
| Right.Hippocampus \| EstimatedTotalIntraCranialVol+CAT12+parent_educ+puberty | -0.0172 | 0.0070 | -2.4684 | 0.0145 | -0.0310 | -0.0035 | TRUE |
| Right.Hippocampus \| age+sex+BrainSegVol+race+parent_educ+puberty | -0.0173 | 0.0077 | -2.2311 | 0.0270 | -0.0326 | -0.0020 | TRUE |
| Right.Hippocampus \| age+sex+BrainSegVol | -0.0173 | 0.0072 | -2.3903 | 0.0178 | -0.0316 | -0.0030 | TRUE |
| Right.Hippocampus \| age+sex+TotalGrayVol+race+puberty | -0.0175 | 0.0078 | -2.2330 | 0.0268 | -0.0329 | -0.0020 | TRUE |
| Right.Hippocampus \| age+sex+BrainSegVol+CAT12+race+parent_educ+puberty | -0.0176 | 0.0078 | -2.2631 | 0.0249 | -0.0329 | -0.0022 | TRUE |
| Right.Hippocampus \| TotalGrayVol+race+puberty | -0.0176 | 0.0079 | -2.2236 | 0.0274 | -0.0332 | -0.0020 | TRUE |
| Right.Hippocampus \| age+sex+TotalGrayVol+CAT12+race+puberty | -0.0176 | 0.0078 | -2.2459 | 0.0260 | -0.0331 | -0.0021 | TRUE |
| Right.Hippocampus \| TotalGrayVol+CAT12+race+puberty | -0.0176 | 0.0079 | -2.2208 | 0.0276 | -0.0332 | -0.0020 | TRUE |
| Right.Hippocampus \| age+sex+CAT12+puberty | -0.0176 | 0.0061 | -2.8759 | 0.0045 | -0.0297 | -0.0055 | TRUE |
| Right.Hippocampus \| parent_educ | -0.0177 | 0.0059 | -3.0044 | 0.0030 | -0.0293 | -0.0061 | TRUE |
| Right.Hippocampus \| CAT12+parent_educ | -0.0178 | 0.0059 | -3.0042 | 0.0030 | -0.0294 | -0.0061 | TRUE |
| Right.Hippocampus \| age+sex+EstimatedTotalIntraCranialVol+puberty | -0.0178 | 0.0067 | -2.6386 | 0.0090 | -0.0310 | -0.0045 | TRUE |
| Right.Hippocampus \| age+sex+BrainSegVol+CAT12 | -0.0178 | 0.0073 | -2.4523 | 0.0151 | -0.0321 | -0.0035 | TRUE |
| Right.Hippocampus \| age+sex+EstimatedTotalIntraCranialVol+CAT12+puberty | -0.0178 | 0.0067 | -2.6435 | 0.0089 | -0.0311 | -0.0045 | TRUE |
| Right.Hippocampus \| age+sex+BrainSegVol+race+puberty | -0.0179 | 0.0077 | -2.3230 | 0.0213 | -0.0331 | -0.0027 | TRUE |
| Right.Hippocampus \| TotalGrayVol+parent_educ+puberty | -0.0179 | 0.0077 | -2.3296 | 0.0209 | -0.0331 | -0.0027 | TRUE |
| Right.Hippocampus \| age+sex+puberty | -0.0179 | 0.0061 | -2.9296 | 0.0038 | -0.0300 | -0.0059 | TRUE |
| Right.Hippocampus \| TotalGrayVol+CAT12+parent_educ+puberty | -0.0180 | 0.0077 | -2.3328 | 0.0208 | -0.0332 | -0.0028 | TRUE |
| Left.Hippocampus \| age+sex+BrainSegVol+puberty | -0.0180 | 0.0082 | -2.2081 | 0.0284 | -0.0342 | -0.0019 | TRUE |
| Right.Hippocampus \| age+sex+BrainSegVol+CAT12+race+puberty | -0.0181 | 0.0077 | -2.3519 | 0.0198 | -0.0333 | -0.0029 | TRUE |
| Right.Hippocampus \| race | -0.0182 | 0.0060 | -3.0457 | 0.0027 | -0.0300 | -0.0064 | TRUE |
| Right.Hippocampus \| CAT12+race | -0.0182 | 0.0060 | -3.0404 | 0.0027 | -0.0301 | -0.0064 | TRUE |
| Left.Hippocampus \| BrainSegVol+puberty | -0.0183 | 0.0082 | -2.2191 | 0.0277 | -0.0346 | -0.0020 | TRUE |
| Right.Hippocampus \| age+sex+TotalGrayVol+parent_educ+puberty | -0.0183 | 0.0077 | -2.3938 | 0.0177 | -0.0334 | -0.0032 | TRUE |
| Left.Hippocampus \| race+parent_educ+puberty | -0.0184 | 0.0070 | -2.6358 | 0.0091 | -0.0323 | -0.0046 | TRUE |
| Left.Hippocampus \| race | -0.0185 | 0.0067 | -2.7505 | 0.0066 | -0.0317 | -0.0052 | TRUE |
| Left.Hippocampus \| CAT12+race+parent_educ+puberty | -0.0185 | 0.0070 | -2.6374 | 0.0091 | -0.0324 | -0.0047 | TRUE |
| Left.Hippocampus \| CAT12+race | -0.0185 | 0.0067 | -2.7510 | 0.0065 | -0.0318 | -0.0052 | TRUE |
| Right.Hippocampus \| age+sex+TotalGrayVol+CAT12+parent_educ+puberty | -0.0185 | 0.0077 | -2.4188 | 0.0166 | -0.0337 | -0.0034 | TRUE |
| Left.Hippocampus \| BrainSegVol+CAT12+puberty | -0.0186 | 0.0083 | -2.2396 | 0.0263 | -0.0349 | -0.0022 | TRUE |
| Right.Hippocampus \| BrainSegVol+race+parent_educ+puberty | -0.0186 | 0.0078 | -2.3777 | 0.0185 | -0.0340 | -0.0032 | TRUE |
| Left.Hippocampus \| age+sex+BrainSegVol+CAT12+puberty | -0.0187 | 0.0082 | -2.2775 | 0.0239 | -0.0348 | -0.0025 | TRUE |
| Right.Hippocampus \| BrainSegVol+CAT12+race+parent_educ+puberty | -0.0187 | 0.0078 | -2.3812 | 0.0183 | -0.0342 | -0.0032 | TRUE |
| Right.Hippocampus \| EstimatedTotalIntraCranialVol+puberty | -0.0187 | 0.0067 | -2.7950 | 0.0057 | -0.0319 | -0.0055 | TRUE |
| Right.Hippocampus \| EstimatedTotalIntraCranialVol+CAT12+puberty | -0.0188 | 0.0067 | -2.7986 | 0.0057 | -0.0320 | -0.0055 | TRUE |
| Right.Hippocampus \| age+sex+BrainSegVol+parent_educ+puberty | -0.0190 | 0.0075 | -2.5239 | 0.0125 | -0.0339 | -0.0042 | TRUE |
| Left.Hippocampus \| parent_educ+puberty | -0.0191 | 0.0067 | -2.8336 | 0.0051 | -0.0324 | -0.0058 | TRUE |
| Right.Hippocampus \| TotalGrayVol+puberty | -0.0192 | 0.0074 | -2.5889 | 0.0104 | -0.0338 | -0.0046 | TRUE |
| Left.Hippocampus \| CAT12+parent_educ+puberty | -0.0192 | 0.0068 | -2.8400 | 0.0050 | -0.0325 | -0.0059 | TRUE |
| Right.Hippocampus \| TotalGrayVol+CAT12+puberty | -0.0192 | 0.0074 | -2.5929 | 0.0103 | -0.0339 | -0.0046 | TRUE |
| Right.Hippocampus \| BrainSegVol+race+puberty | -0.0193 | 0.0078 | -2.4760 | 0.0142 | -0.0346 | -0.0039 | TRUE |
| Right.Hippocampus \| BrainSegVol+CAT12+race+puberty | -0.0193 | 0.0078 | -2.4740 | 0.0143 | -0.0348 | -0.0039 | TRUE |
| Right.Hippocampus \| race+parent_educ+puberty | -0.0194 | 0.0061 | -3.2030 | 0.0016 | -0.0313 | -0.0074 | TRUE |
| Right.Hippocampus \| CAT12+race+parent_educ+puberty | -0.0194 | 0.0061 | -3.1982 | 0.0016 | -0.0314 | -0.0074 | TRUE |
| Right.Hippocampus \| age+sex+BrainSegVol+CAT12+parent_educ+puberty | -0.0194 | 0.0076 | -2.5684 | 0.0110 | -0.0343 | -0.0045 | TRUE |
| Right.Hippocampus \| BrainSegVol+parent_educ+puberty | -0.0196 | 0.0076 | -2.5847 | 0.0105 | -0.0346 | -0.0046 | TRUE |
| Right.Hippocampus \| BrainSegVol+CAT12+parent_educ+puberty | -0.0198 | 0.0076 | -2.5941 | 0.0103 | -0.0349 | -0.0047 | TRUE |
| Right.Hippocampus \| age+sex+TotalGrayVol+puberty | -0.0198 | 0.0074 | -2.6840 | 0.0079 | -0.0344 | -0.0053 | TRUE |
| Right.Hippocampus \| age+sex+TotalGrayVol+CAT12+puberty | -0.0200 | 0.0074 | -2.7128 | 0.0073 | -0.0346 | -0.0055 | TRUE |
| Right.Hippocampus \| CAT12 | -0.0202 | 0.0056 | -3.6062 | 0.0004 | -0.0312 | -0.0091 | TRUE |
| Left.Hippocampus \| unadjusted | -0.0202 | 0.0063 | -3.2249 | 0.0015 | -0.0325 | -0.0078 | TRUE |
| Right.Hippocampus \| unadjusted | -0.0202 | 0.0056 | -3.6167 | 0.0004 | -0.0312 | -0.0092 | TRUE |
| Left.Hippocampus \| CAT12 | -0.0202 | 0.0063 | -3.2202 | 0.0015 | -0.0326 | -0.0078 | TRUE |
| Right.Hippocampus \| parent_educ+puberty | -0.0203 | 0.0058 | -3.4791 | 0.0006 | -0.0318 | -0.0088 | TRUE |
| Right.Hippocampus \| CAT12+parent_educ+puberty | -0.0203 | 0.0058 | -3.4768 | 0.0006 | -0.0319 | -0.0088 | TRUE |
| Left.Hippocampus \| race+puberty | -0.0206 | 0.0066 | -3.1129 | 0.0022 | -0.0337 | -0.0076 | TRUE |
| Left.Hippocampus \| CAT12+race+puberty | -0.0207 | 0.0067 | -3.1131 | 0.0022 | -0.0339 | -0.0076 | TRUE |
| Right.Hippocampus \| race+puberty | -0.0208 | 0.0059 | -3.5175 | 0.0006 | -0.0324 | -0.0091 | TRUE |
| Right.Hippocampus \| CAT12+race+puberty | -0.0208 | 0.0059 | -3.5110 | 0.0006 | -0.0325 | -0.0091 | TRUE |
| Right.Hippocampus \| age+sex+BrainSegVol+puberty | -0.0209 | 0.0073 | -2.8671 | 0.0046 | -0.0352 | -0.0065 | TRUE |
| Right.Hippocampus \| BrainSegVol+puberty | -0.0211 | 0.0073 | -2.8803 | 0.0044 | -0.0356 | -0.0067 | TRUE |
| Right.Hippocampus \| age+sex+BrainSegVol+CAT12+puberty | -0.0213 | 0.0073 | -2.9190 | 0.0039 | -0.0356 | -0.0069 | TRUE |
| Right.Hippocampus \| BrainSegVol+CAT12+puberty | -0.0213 | 0.0074 | -2.8937 | 0.0043 | -0.0358 | -0.0068 | TRUE |
| Left.Hippocampus \| puberty | -0.0221 | 0.0062 | -3.5817 | 0.0004 | -0.0343 | -0.0099 | TRUE |
| Left.Hippocampus \| CAT12+puberty | -0.0222 | 0.0062 | -3.5793 | 0.0004 | -0.0344 | -0.0099 | TRUE |
| Right.Hippocampus \| CAT12+puberty | -0.0225 | 0.0055 | -4.0881 | 0.0001 | -0.0334 | -0.0117 | TRUE |
| Right.Hippocampus \| puberty | -0.0225 | 0.0055 | -4.0978 | 0.0001 | -0.0334 | -0.0117 | TRUE |

**Table S2. Multiverse results: amygdala volume → stress severity (stressth)**

*384 model specifications produced a valid fit. β = unstandardized OLS coefficient; SE = standard error; CI = 95% confidence interval. Values rounded to four decimal places.*

| **model_label** | **estimate** | **SE** | **t** | **p** | **CI low** | **CI high** | **significant** |
| --- | --- | --- | --- | --- | --- | --- | --- |
| Total.Amygdala \| age+sex+CAT12+race+parent_educ | -0.0132 | 0.0080 | -1.638 | 0.1032 | -0.0291 | 0.0027 | FALSE |
| Total.Amygdala \| age+sex+race+parent_educ | -0.0139 | 0.0080 | -1.7411 | 0.0834 | -0.0300 | 0.0019 | FALSE |
| Right.Amygdala \| age+sex+CAT12+race+parent_educ | -0.0140 | 0.0150 | -0.9348 | 0.3512 | -0.0437 | 0.0156 | FALSE |
| Total.Amygdala \| age+sex+CAT12+race | -0.0146 | 0.0077 | -1.8919 | 0.0601 | -0.0299 | 0.0006 | FALSE |
| Total.Amygdala \| age+sex+CAT12+race+parent_educ+puberty | -0.0155 | 0.0081 | -1.9279 | 0.0555 | -0.0314 | 0.0004 | FALSE |
| Right.Amygdala \| age+sex+race+parent_educ | -0.0155 | 0.0149 | -1.0437 | 0.2980 | -0.0449 | 0.0138 | FALSE |
| Total.Amygdala \| age+sex+EstimatedTotalIntraCranialVol+CAT12+race+parent_educ | -0.0159 | 0.0090 | -1.7773 | 0.0773 | -0.0336 | 0.0018 | FALSE |
| Total.Amygdala \| age+sex+EstimatedTotalIntraCranialVol+race+parent_educ | -0.0164 | 0.0089 | -1.8387 | 0.0677 | -0.0340 | 0.0012 | FALSE |
| Right.Amygdala \| age+sex+EstimatedTotalIntraCranialVol+CAT12+race+parent_educ | -0.0165 | 0.0167 | -0.9916 | 0.3227 | -0.0494 | 0.0164 | FALSE |
| Total.Amygdala \| age+sex+EstimatedTotalIntraCranialVol+CAT12+race+parent_educ+puberty | -0.0174 | 0.0089 | -1.9460 | 0.0533 | -0.0350 | 0.0002 | FALSE |
| Right.Amygdala \| age+sex+EstimatedTotalIntraCranialVol+race+parent_educ | -0.0176 | 0.0166 | -1.0569 | 0.2920 | -0.0503 | 0.0152 | FALSE |
| Right.Amygdala \| age+sex+CAT12+race | -0.0176 | 0.0146 | -1.2044 | 0.2300 | -0.0465 | 0.0113 | FALSE |
| Right.Amygdala \| age+sex+CAT12+race+parent_educ+puberty | -0.0182 | 0.0151 | -1.2114 | 0.2274 | -0.0480 | 0.0115 | FALSE |
| Right.Amygdala \| age+sex+race | -0.0191 | 0.0145 | -1.3164 | 0.1897 | -0.0476 | 0.0095 | FALSE |
| Right.Amygdala \| age+sex+EstimatedTotalIntraCranialVol+CAT12+race+parent_educ+puberty | -0.0192 | 0.0166 | -1.1547 | 0.2498 | -0.0520 | 0.0136 | FALSE |
| Right.Amygdala \| age+sex+race+parent_educ+puberty | -0.0196 | 0.0149 | -1.3137 | 0.1907 | -0.0490 | 0.0098 | FALSE |
| Right.Amygdala \| age+sex+EstimatedTotalIntraCranialVol+race+parent_educ+puberty | -0.0201 | 0.0165 | -1.2156 | 0.2258 | -0.0528 | 0.0125 | FALSE |
| Right.Amygdala \| age+sex+EstimatedTotalIntraCranialVol+CAT12+race | -0.0202 | 0.0163 | -1.2334 | 0.2191 | -0.0524 | 0.0121 | FALSE |
| Right.Amygdala \| age+sex+CAT12+parent_educ | -0.0202 | 0.0147 | -1.3750 | 0.1708 | -0.0491 | 0.0088 | FALSE |
| Right.Amygdala \| age+sex+TotalGrayVol+CAT12+race+parent_educ | -0.0203 | 0.0178 | -1.1408 | 0.2555 | -0.0554 | 0.0148 | FALSE |
| Right.Amygdala \| age+sex+EstimatedTotalIntraCranialVol+race | -0.0211 | 0.0163 | -1.2985 | 0.1958 | -0.0532 | 0.0110 | FALSE |
| Right.Amygdala \| age+sex+TotalGrayVol+race+parent_educ | -0.0213 | 0.0177 | -1.2021 | 0.2309 | -0.0563 | 0.0137 | FALSE |
| Right.Amygdala \| age+sex+EstimatedTotalIntraCranialVol+CAT12+parent_educ | -0.0214 | 0.0164 | -1.3053 | 0.1934 | -0.0538 | 0.0110 | FALSE |
| Right.Amygdala \| age+sex+parent_educ | -0.0215 | 0.0145 | -1.4780 | 0.1411 | -0.0502 | 0.0072 | FALSE |
| Right.Amygdala \| age+sex+CAT12+race+puberty | -0.0218 | 0.0147 | -1.4838 | 0.1396 | -0.0507 | 0.0072 | FALSE |
| Right.Amygdala \| age+sex+EstimatedTotalIntraCranialVol+parent_educ | -0.0223 | 0.0164 | -1.3610 | 0.1752 | -0.0545 | 0.0100 | FALSE |
| Right.Amygdala \| age+sex+EstimatedTotalIntraCranialVol+CAT12+race+puberty | -0.0228 | 0.0163 | -1.3992 | 0.1635 | -0.0549 | 0.0094 | FALSE |
| Right.Amygdala \| age+sex+TotalGrayVol+CAT12+race+parent_educ+puberty | -0.0228 | 0.0177 | -1.2859 | 0.2002 | -0.0578 | 0.0122 | FALSE |
| Right.Amygdala \| age+sex+race+puberty | -0.0230 | 0.0145 | -1.5884 | 0.1140 | -0.0516 | 0.0056 | FALSE |
| Right.Amygdala \| age+sex+BrainSegVol+CAT12+race+parent_educ | -0.0230 | 0.0180 | -1.2807 | 0.2020 | -0.0585 | 0.0125 | FALSE |
| Right.Amygdala \| age+sex+EstimatedTotalIntraCranialVol+race+puberty | -0.0236 | 0.0162 | -1.4595 | 0.1462 | -0.0556 | 0.0083 | FALSE |
| Right.Amygdala \| TotalGrayVol+CAT12+race+parent_educ | -0.0237 | 0.0180 | -1.3139 | 0.1906 | -0.0592 | 0.0119 | FALSE |
| Right.Amygdala \| age+sex+BrainSegVol+race+parent_educ | -0.0237 | 0.0180 | -1.3189 | 0.1889 | -0.0591 | 0.0118 | FALSE |
| Right.Amygdala \| age+sex+TotalGrayVol+race+parent_educ+puberty | -0.0237 | 0.0177 | -1.3446 | 0.1805 | -0.0586 | 0.0111 | FALSE |
| Right.Amygdala \| age+sex+EstimatedTotalIntraCranialVol+CAT12+parent_educ+puberty | -0.0238 | 0.0163 | -1.4573 | 0.1468 | -0.0560 | 0.0084 | FALSE |
| Right.Amygdala \| TotalGrayVol+race+parent_educ | -0.0238 | 0.0179 | -1.3279 | 0.1859 | -0.0592 | 0.0116 | FALSE |
| Right.Amygdala \| age+sex+CAT12+parent_educ+puberty | -0.0239 | 0.0146 | -1.6340 | 0.1040 | -0.0528 | 0.0050 | FALSE |
| Right.Amygdala \| age+sex+TotalGrayVol+CAT12+race | -0.0239 | 0.0175 | -1.3695 | 0.1726 | -0.0584 | 0.0106 | FALSE |
| Right.Amygdala \| age+sex+TotalGrayVol+CAT12+parent_educ | -0.0241 | 0.0176 | -1.3728 | 0.1715 | -0.0588 | 0.0105 | FALSE |
| Right.Amygdala \| age+sex+EstimatedTotalIntraCranialVol+parent_educ+puberty | -0.0245 | 0.0162 | -1.5084 | 0.1332 | -0.0566 | 0.0076 | FALSE |
| Right.Amygdala \| EstimatedTotalIntraCranialVol+CAT12+race+parent_educ | -0.0248 | 0.0166 | -1.4901 | 0.1380 | -0.0576 | 0.0080 | FALSE |
| Right.Amygdala \| age+sex+TotalGrayVol+race | -0.0249 | 0.0174 | -1.4273 | 0.1553 | -0.0592 | 0.0095 | FALSE |
| Right.Amygdala \| EstimatedTotalIntraCranialVol+race+parent_educ | -0.0249 | 0.0166 | -1.4999 | 0.1354 | -0.0576 | 0.0079 | FALSE |
| Right.Amygdala \| age+sex+TotalGrayVol+parent_educ | -0.0249 | 0.0175 | -1.4244 | 0.1560 | -0.0595 | 0.0096 | FALSE |
| Right.Amygdala \| age+sex+parent_educ+puberty | -0.0251 | 0.0145 | -1.7289 | 0.0855 | -0.0537 | 0.0035 | FALSE |
| Right.Amygdala \| age+sex+EstimatedTotalIntraCranialVol+CAT12 | -0.0251 | 0.0157 | -1.5951 | 0.1123 | -0.0561 | 0.0059 | FALSE |
| Right.Amygdala \| age+sex+CAT12 | -0.0252 | 0.0140 | -1.8039 | 0.0728 | -0.0528 | 0.0024 | FALSE |
| Right.Amygdala \| age+sex+BrainSegVol+CAT12+race+parent_educ+puberty | -0.0256 | 0.0180 | -1.4252 | 0.1559 | -0.0610 | 0.0098 | FALSE |
| Right.Amygdala \| age+sex+EstimatedTotalIntraCranialVol | -0.0260 | 0.0157 | -1.6558 | 0.0994 | -0.0569 | 0.0050 | FALSE |
| Right.Amygdala \| age+sex+BrainSegVol+race+parent_educ+puberty | -0.0262 | 0.0179 | -1.4648 | 0.1448 | -0.0616 | 0.0091 | FALSE |
| Right.Amygdala \| TotalGrayVol+CAT12+parent_educ | -0.0264 | 0.0177 | -1.4890 | 0.1382 | -0.0613 | 0.0086 | FALSE |
| Right.Amygdala \| TotalGrayVol+CAT12+race+parent_educ+puberty | -0.0264 | 0.0178 | -1.4810 | 0.1404 | -0.0615 | 0.0088 | FALSE |
| Right.Amygdala \| age+sex+TotalGrayVol+CAT12+race+puberty | -0.0264 | 0.0174 | -1.5166 | 0.1312 | -0.0608 | 0.0080 | FALSE |
| Right.Amygdala \| EstimatedTotalIntraCranialVol+CAT12+race+parent_educ+puberty | -0.0265 | 0.0164 | -1.6121 | 0.1088 | -0.0589 | 0.0059 | FALSE |
| Right.Amygdala \| TotalGrayVol+race+parent_educ+puberty | -0.0265 | 0.0177 | -1.4944 | 0.1369 | -0.0614 | 0.0085 | FALSE |
| Right.Amygdala \| EstimatedTotalIntraCranialVol+race+parent_educ+puberty | -0.0265 | 0.0164 | -1.6214 | 0.1067 | -0.0588 | 0.0058 | FALSE |
| Right.Amygdala \| TotalGrayVol+parent_educ | -0.0265 | 0.0176 | -1.5054 | 0.1339 | -0.0613 | 0.0082 | FALSE |
| Right.Amygdala \| age+sex+TotalGrayVol+CAT12 | -0.0266 | 0.0168 | -1.5847 | 0.1147 | -0.0597 | 0.0065 | FALSE |
| Right.Amygdala \| age+sex | -0.0267 | 0.0138 | -1.9266 | 0.0555 | -0.0540 | 0.0006 | FALSE |
| Right.Amygdala \| age+sex+TotalGrayVol+CAT12+parent_educ+puberty | -0.0268 | 0.0175 | -1.5328 | 0.1271 | -0.0613 | 0.0077 | FALSE |
| Right.Amygdala \| age+sex+BrainSegVol+CAT12+race | -0.0268 | 0.0176 | -1.5222 | 0.1297 | -0.0616 | 0.0080 | FALSE |
| Right.Amygdala \| age+sex+TotalGrayVol+race+puberty | -0.0273 | 0.0174 | -1.5716 | 0.1178 | -0.0615 | 0.0070 | FALSE |
| Right.Amygdala \| TotalGrayVol+CAT12+race | -0.0273 | 0.0178 | -1.5387 | 0.1257 | -0.0624 | 0.0077 | FALSE |
| Right.Amygdala \| TotalGrayVol+race | -0.0274 | 0.0177 | -1.5469 | 0.1237 | -0.0623 | 0.0075 | FALSE |
| Right.Amygdala \| age+sex+TotalGrayVol | -0.0274 | 0.0167 | -1.6362 | 0.1034 | -0.0604 | 0.0056 | FALSE |
| Right.Amygdala \| age+sex+BrainSegVol+race | -0.0275 | 0.0176 | -1.5596 | 0.1206 | -0.0622 | 0.0073 | FALSE |
| Right.Amygdala \| age+sex+EstimatedTotalIntraCranialVol+CAT12+puberty | -0.0275 | 0.0157 | -1.7580 | 0.0804 | -0.0584 | 0.0034 | FALSE |
| Right.Amygdala \| age+sex+TotalGrayVol+parent_educ+puberty | -0.0276 | 0.0174 | -1.5824 | 0.1153 | -0.0620 | 0.0068 | FALSE |
| Right.Amygdala \| age+sex+BrainSegVol+CAT12+parent_educ | -0.0276 | 0.0177 | -1.5575 | 0.1211 | -0.0625 | 0.0074 | FALSE |
| Right.Amygdala \| EstimatedTotalIntraCranialVol+CAT12+parent_educ | -0.0277 | 0.0164 | -1.6939 | 0.0920 | -0.0600 | 0.0046 | FALSE |
| Right.Amygdala \| EstimatedTotalIntraCranialVol+parent_educ | -0.0278 | 0.0163 | -1.7057 | 0.0898 | -0.0600 | 0.0044 | FALSE |
| Right.Amygdala \| age+sex+BrainSegVol+parent_educ | -0.0281 | 0.0177 | -1.5876 | 0.1141 | -0.0629 | 0.0068 | FALSE |
| Right.Amygdala \| age+sex+EstimatedTotalIntraCranialVol+puberty | -0.0283 | 0.0156 | -1.8167 | 0.0708 | -0.0591 | 0.0024 | FALSE |
| Right.Amygdala \| TotalGrayVol+CAT12 | -0.0287 | 0.0169 | -1.6961 | 0.0915 | -0.0620 | 0.0047 | FALSE |
| Right.Amygdala \| EstimatedTotalIntraCranialVol+CAT12+race | -0.0287 | 0.0164 | -1.7550 | 0.0810 | -0.0610 | 0.0036 | FALSE |
| Right.Amygdala \| EstimatedTotalIntraCranialVol+race | -0.0287 | 0.0163 | -1.7615 | 0.0798 | -0.0609 | 0.0035 | FALSE |
| Right.Amygdala \| BrainSegVol+CAT12+race+parent_educ | -0.0288 | 0.0183 | -1.5736 | 0.1174 | -0.0648 | 0.0073 | FALSE |
| Right.Amygdala \| BrainSegVol+race+parent_educ | -0.0288 | 0.0182 | -1.5801 | 0.1159 | -0.0647 | 0.0072 | FALSE |
| Right.Amygdala \| TotalGrayVol | -0.0289 | 0.0168 | -1.7124 | 0.0884 | -0.0621 | 0.0044 | FALSE |
| Right.Amygdala \| EstimatedTotalIntraCranialVol+CAT12+parent_educ+puberty | -0.0291 | 0.0161 | -1.8046 | 0.0728 | -0.0609 | 0.0027 | FALSE |
| Right.Amygdala \| EstimatedTotalIntraCranialVol+parent_educ+puberty | -0.0292 | 0.0161 | -1.8158 | 0.0710 | -0.0609 | 0.0025 | FALSE |
| Right.Amygdala \| TotalGrayVol+CAT12+parent_educ+puberty | -0.0292 | 0.0175 | -1.6682 | 0.0970 | -0.0638 | 0.0053 | FALSE |
| Right.Amygdala \| age+sex+TotalGrayVol+CAT12+puberty | -0.0293 | 0.0167 | -1.7521 | 0.0814 | -0.0623 | 0.0037 | FALSE |
| Right.Amygdala \| TotalGrayVol+parent_educ+puberty | -0.0294 | 0.0174 | -1.6842 | 0.0938 | -0.0638 | 0.0050 | FALSE |
| Right.Amygdala \| age+sex+BrainSegVol+CAT12+race+puberty | -0.0294 | 0.0176 | -1.6718 | 0.0964 | -0.0641 | 0.0053 | FALSE |
| Right.Amygdala \| TotalGrayVol+CAT12+race+puberty | -0.0300 | 0.0175 | -1.7105 | 0.0889 | -0.0646 | 0.0046 | FALSE |
| Right.Amygdala \| TotalGrayVol+race+puberty | -0.0300 | 0.0174 | -1.7190 | 0.0874 | -0.0644 | 0.0044 | FALSE |
| Right.Amygdala \| age+sex+BrainSegVol+race+puberty | -0.0300 | 0.0176 | -1.7101 | 0.0890 | -0.0646 | 0.0046 | FALSE |
| Right.Amygdala \| age+sex+TotalGrayVol+puberty | -0.0301 | 0.0167 | -1.8035 | 0.0729 | -0.0629 | 0.0028 | FALSE |
| Right.Amygdala \| EstimatedTotalIntraCranialVol+CAT12+race+puberty | -0.0302 | 0.0161 | -1.8731 | 0.0627 | -0.0619 | 0.0016 | FALSE |
| Right.Amygdala \| EstimatedTotalIntraCranialVol+race+puberty | -0.0302 | 0.0160 | -1.8798 | 0.0618 | -0.0618 | 0.0015 | FALSE |
| Right.Amygdala \| age+sex+BrainSegVol+CAT12+parent_educ+puberty | -0.0303 | 0.0176 | -1.7183 | 0.0875 | -0.0651 | 0.0045 | FALSE |
| Right.Amygdala \| age+sex+BrainSegVol+CAT12 | -0.0306 | 0.0169 | -1.8063 | 0.0725 | -0.0639 | 0.0028 | FALSE |
| Right.Amygdala \| age+sex+BrainSegVol+parent_educ+puberty | -0.0308 | 0.0176 | -1.7504 | 0.0817 | -0.0655 | 0.0039 | FALSE |
| Right.Amygdala \| EstimatedTotalIntraCranialVol+CAT12 | -0.0309 | 0.0157 | -1.9683 | 0.0505 | -0.0618 | 0.0001 | FALSE |
| Right.Amygdala \| age+sex+BrainSegVol | -0.0310 | 0.0169 | -1.8350 | 0.0680 | -0.0644 | 0.0023 | FALSE |
| Right.Amygdala \| BrainSegVol+CAT12+race+parent_educ+puberty | -0.0311 | 0.0181 | -1.7204 | 0.0871 | -0.0667 | 0.0046 | FALSE |
| Right.Amygdala \| BrainSegVol+race+parent_educ+puberty | -0.0311 | 0.0180 | -1.7272 | 0.0859 | -0.0666 | 0.0044 | FALSE |
| Right.Amygdala \| TotalGrayVol+CAT12+puberty | -0.0315 | 0.0167 | -1.8863 | 0.0608 | -0.0645 | 0.0014 | FALSE |
| Right.Amygdala \| TotalGrayVol+puberty | -0.0317 | 0.0167 | -1.9062 | 0.0581 | -0.0646 | 0.0011 | FALSE |
| Right.Amygdala \| BrainSegVol+CAT12+parent_educ | -0.0319 | 0.0179 | -1.7834 | 0.0762 | -0.0673 | 0.0034 | FALSE |
| Right.Amygdala \| BrainSegVol+parent_educ | -0.0320 | 0.0179 | -1.7905 | 0.0750 | -0.0672 | 0.0033 | FALSE |
| Right.Amygdala \| BrainSegVol+race | -0.0327 | 0.0179 | -1.8229 | 0.0700 | -0.0681 | 0.0027 | FALSE |
| Right.Amygdala \| BrainSegVol+CAT12+race | -0.0327 | 0.0180 | -1.8177 | 0.0708 | -0.0682 | 0.0028 | FALSE |
| Right.Amygdala \| BrainSegVol+CAT12+parent_educ+puberty | -0.0344 | 0.0177 | -1.9452 | 0.0533 | -0.0694 | 0.0005 | FALSE |
| Right.Amygdala \| BrainSegVol+parent_educ+puberty | -0.0345 | 0.0177 | -1.9530 | 0.0523 | -0.0693 | 0.0004 | FALSE |
| Right.Amygdala \| BrainSegVol+CAT12+race+puberty | -0.0350 | 0.0177 | -1.9722 | 0.0501 | -0.0700 | 0.0000 | FALSE |
| Total.Amygdala \| age+sex+race | -0.0153 | 0.0077 | -2.0004 | 0.0470 | -0.0304 | -0.0002 | TRUE |
| Total.Amygdala \| age+sex+race+parent_educ+puberty | -0.0161 | 0.0080 | -2.0235 | 0.0446 | -0.0318 | -0.0004 | TRUE |
| Total.Amygdala \| age+sex+CAT12+parent_educ | -0.0163 | 0.0079 | -2.0778 | 0.0391 | -0.0318 | -0.0008 | TRUE |
| Total.Amygdala \| age+sex+CAT12+race+puberty | -0.0169 | 0.0077 | -2.1792 | 0.0306 | -0.0322 | -0.0016 | TRUE |
| Total.Amygdala \| age+sex+parent_educ | -0.0170 | 0.0078 | -2.1743 | 0.0310 | -0.0323 | -0.0016 | TRUE |
| Total.Amygdala \| age+sex+race+puberty | -0.0174 | 0.0077 | -2.2791 | 0.0239 | -0.0325 | -0.0023 | TRUE |
| Total.Amygdala \| age+sex+EstimatedTotalIntraCranialVol+CAT12+race | -0.0175 | 0.0087 | -2.0108 | 0.0459 | -0.0347 | -0.0003 | TRUE |
| Total.Amygdala \| age+sex+EstimatedTotalIntraCranialVol+race+parent_educ+puberty | -0.0178 | 0.0089 | -2.0027 | 0.0468 | -0.0353 | -0.0003 | TRUE |
| Total.Amygdala \| age+sex+EstimatedTotalIntraCranialVol+race | -0.0180 | 0.0087 | -2.0748 | 0.0394 | -0.0351 | -0.0009 | TRUE |
| Total.Amygdala \| age+sex+CAT12+parent_educ+puberty | -0.0184 | 0.0078 | -2.3472 | 0.0200 | -0.0339 | -0.0029 | TRUE |
| Total.Amygdala \| age+sex+EstimatedTotalIntraCranialVol+CAT12+parent_educ | -0.0185 | 0.0088 | -2.0933 | 0.0377 | -0.0359 | -0.0011 | TRUE |
| Total.Amygdala \| age+sex+EstimatedTotalIntraCranialVol+CAT12+race+puberty | -0.0188 | 0.0087 | -2.1749 | 0.0310 | -0.0359 | -0.0017 | TRUE |
| Total.Amygdala \| age+sex+CAT12 | -0.0189 | 0.0074 | -2.5463 | 0.0117 | -0.0335 | -0.0043 | TRUE |
| Total.Amygdala \| age+sex+EstimatedTotalIntraCranialVol+parent_educ | -0.0189 | 0.0088 | -2.1440 | 0.0334 | -0.0363 | -0.0015 | TRUE |
| Total.Amygdala \| age+sex+parent_educ+puberty | -0.0189 | 0.0078 | -2.4352 | 0.0158 | -0.0342 | -0.0036 | TRUE |
| Total.Amygdala \| age+sex+EstimatedTotalIntraCranialVol+race+puberty | -0.0192 | 0.0086 | -2.2332 | 0.0268 | -0.0362 | -0.0022 | TRUE |
| Total.Amygdala \| age+sex+TotalGrayVol+CAT12+race+parent_educ | -0.0193 | 0.0096 | -2.0072 | 0.0463 | -0.0384 | -0.0003 | TRUE |
| Total.Amygdala \| EstimatedTotalIntraCranialVol+CAT12+race+parent_educ | -0.0194 | 0.0090 | -2.1670 | 0.0316 | -0.0371 | -0.0017 | TRUE |
| Total.Amygdala \| EstimatedTotalIntraCranialVol+race+parent_educ | -0.0195 | 0.0089 | -2.1784 | 0.0307 | -0.0371 | -0.0018 | TRUE |
| Total.Amygdala \| age+sex | -0.0196 | 0.0073 | -2.6739 | 0.0081 | -0.0340 | -0.0051 | TRUE |
| Total.Amygdala \| age+sex+EstimatedTotalIntraCranialVol+CAT12+parent_educ+puberty | -0.0197 | 0.0088 | -2.2490 | 0.0257 | -0.0370 | -0.0024 | TRUE |
| Total.Amygdala \| age+sex+TotalGrayVol+race+parent_educ | -0.0198 | 0.0096 | -2.0639 | 0.0405 | -0.0388 | -0.0009 | TRUE |
| Total.Amygdala \| TotalGrayVol+CAT12+race+parent_educ | -0.0199 | 0.0098 | -2.0403 | 0.0428 | -0.0392 | -0.0007 | TRUE |
| Total.Amygdala \| TotalGrayVol+race+parent_educ | -0.0200 | 0.0097 | -2.0556 | 0.0413 | -0.0392 | -0.0008 | TRUE |
| Total.Amygdala \| age+sex+EstimatedTotalIntraCranialVol+parent_educ+puberty | -0.0201 | 0.0087 | -2.2953 | 0.0229 | -0.0373 | -0.0028 | TRUE |
| Total.Amygdala \| EstimatedTotalIntraCranialVol+CAT12+race+parent_educ+puberty | -0.0204 | 0.0088 | -2.3153 | 0.0218 | -0.0379 | -0.0030 | TRUE |
| Total.Amygdala \| EstimatedTotalIntraCranialVol+race+parent_educ+puberty | -0.0205 | 0.0088 | -2.3261 | 0.0212 | -0.0378 | -0.0031 | TRUE |
| Total.Amygdala \| age+sex+EstimatedTotalIntraCranialVol+CAT12 | -0.0205 | 0.0084 | -2.4322 | 0.0159 | -0.0371 | -0.0039 | TRUE |
| Total.Amygdala \| age+sex+TotalGrayVol+CAT12+race+parent_educ+puberty | -0.0207 | 0.0096 | -2.1550 | 0.0326 | -0.0396 | -0.0017 | TRUE |
| Total.Amygdala \| EstimatedTotalIntraCranialVol+CAT12+parent_educ | -0.0209 | 0.0088 | -2.3695 | 0.0188 | -0.0383 | -0.0035 | TRUE |
| Total.Amygdala \| age+sex+CAT12+puberty | -0.0209 | 0.0074 | -2.8306 | 0.0051 | -0.0355 | -0.0063 | TRUE |
| Total.Amygdala \| age+sex+EstimatedTotalIntraCranialVol | -0.0210 | 0.0084 | -2.5013 | 0.0132 | -0.0375 | -0.0044 | TRUE |
| Total.Amygdala \| EstimatedTotalIntraCranialVol+parent_educ | -0.0210 | 0.0088 | -2.3827 | 0.0182 | -0.0383 | -0.0036 | TRUE |
| Total.Amygdala \| age+sex+TotalGrayVol+CAT12+race | -0.0210 | 0.0094 | -2.2363 | 0.0266 | -0.0396 | -0.0025 | TRUE |
| Total.Amygdala \| age+sex+TotalGrayVol+race+parent_educ+puberty | -0.0211 | 0.0096 | -2.2088 | 0.0285 | -0.0400 | -0.0022 | TRUE |
| Total.Amygdala \| TotalGrayVol+CAT12+parent_educ | -0.0213 | 0.0096 | -2.2136 | 0.0281 | -0.0403 | -0.0023 | TRUE |
| Total.Amygdala \| age+sex+TotalGrayVol+CAT12+parent_educ | -0.0213 | 0.0095 | -2.2371 | 0.0265 | -0.0402 | -0.0025 | TRUE |
| Total.Amygdala \| TotalGrayVol+parent_educ | -0.0214 | 0.0096 | -2.2312 | 0.0269 | -0.0403 | -0.0025 | TRUE |
| Total.Amygdala \| age+sex+BrainSegVol+CAT12+race+parent_educ | -0.0214 | 0.0098 | -2.1843 | 0.0303 | -0.0407 | -0.0021 | TRUE |
| Total.Amygdala \| EstimatedTotalIntraCranialVol+race | -0.0214 | 0.0087 | -2.4644 | 0.0147 | -0.0386 | -0.0043 | TRUE |
| Total.Amygdala \| EstimatedTotalIntraCranialVol+CAT12+race | -0.0214 | 0.0087 | -2.4564 | 0.0150 | -0.0387 | -0.0042 | TRUE |
| Total.Amygdala \| age+sex+TotalGrayVol+race | -0.0215 | 0.0094 | -2.2925 | 0.0230 | -0.0400 | -0.0030 | TRUE |
| Total.Amygdala \| age+sex+puberty | -0.0216 | 0.0073 | -2.9536 | 0.0035 | -0.0360 | -0.0072 | TRUE |
| Total.Amygdala \| TotalGrayVol+CAT12+race+parent_educ+puberty | -0.0216 | 0.0096 | -2.2386 | 0.0265 | -0.0406 | -0.0026 | TRUE |
| Total.Amygdala \| TotalGrayVol+race+parent_educ+puberty | -0.0216 | 0.0096 | -2.2530 | 0.0255 | -0.0406 | -0.0027 | TRUE |
| Total.Amygdala \| age+sex+BrainSegVol+race+parent_educ | -0.0217 | 0.0098 | -2.2159 | 0.0280 | -0.0410 | -0.0024 | TRUE |
| Total.Amygdala \| age+sex+TotalGrayVol+parent_educ | -0.0217 | 0.0095 | -2.2828 | 0.0236 | -0.0405 | -0.0029 | TRUE |
| Total.Amygdala \| age+sex+EstimatedTotalIntraCranialVol+CAT12+puberty | -0.0218 | 0.0084 | -2.6036 | 0.0100 | -0.0383 | -0.0053 | TRUE |
| Total.Amygdala \| EstimatedTotalIntraCranialVol+CAT12+parent_educ+puberty | -0.0218 | 0.0087 | -2.5083 | 0.0130 | -0.0390 | -0.0047 | TRUE |
| Total.Amygdala \| EstimatedTotalIntraCranialVol+parent_educ+puberty | -0.0218 | 0.0087 | -2.5209 | 0.0126 | -0.0389 | -0.0047 | TRUE |
| Total.Amygdala \| CAT12+race+parent_educ | -0.0220 | 0.0071 | -3.0871 | 0.0023 | -0.0360 | -0.0079 | TRUE |
| Total.Amygdala \| TotalGrayVol+race | -0.0220 | 0.0095 | -2.3019 | 0.0225 | -0.0408 | -0.0031 | TRUE |
| Total.Amygdala \| TotalGrayVol+CAT12+race | -0.0220 | 0.0096 | -2.2925 | 0.0230 | -0.0409 | -0.0031 | TRUE |
| Total.Amygdala \| race+parent_educ | -0.0220 | 0.0071 | -3.1032 | 0.0022 | -0.0360 | -0.0080 | TRUE |
| Total.Amygdala \| EstimatedTotalIntraCranialVol+race+puberty | -0.0222 | 0.0086 | -2.5930 | 0.0103 | -0.0390 | -0.0053 | TRUE |
| Total.Amygdala \| EstimatedTotalIntraCranialVol+CAT12+race+puberty | -0.0222 | 0.0086 | -2.5849 | 0.0105 | -0.0391 | -0.0052 | TRUE |
| Total.Amygdala \| age+sex+EstimatedTotalIntraCranialVol+puberty | -0.0222 | 0.0083 | -2.6692 | 0.0083 | -0.0386 | -0.0058 | TRUE |
| Total.Amygdala \| age+sex+TotalGrayVol+CAT12+race+puberty | -0.0223 | 0.0094 | -2.3780 | 0.0185 | -0.0408 | -0.0038 | TRUE |
| Total.Amygdala \| age+sex+TotalGrayVol+race+puberty | -0.0227 | 0.0093 | -2.4305 | 0.0161 | -0.0411 | -0.0043 | TRUE |
| Total.Amygdala \| age+sex+BrainSegVol+CAT12+race+parent_educ+puberty | -0.0227 | 0.0098 | -2.3300 | 0.0210 | -0.0420 | -0.0035 | TRUE |
| Total.Amygdala \| age+sex+TotalGrayVol+CAT12+parent_educ+puberty | -0.0228 | 0.0095 | -2.4007 | 0.0174 | -0.0415 | -0.0041 | TRUE |
| Total.Amygdala \| age+sex+TotalGrayVol+CAT12 | -0.0228 | 0.0091 | -2.5005 | 0.0132 | -0.0407 | -0.0048 | TRUE |
| Total.Amygdala \| EstimatedTotalIntraCranialVol+CAT12 | -0.0229 | 0.0084 | -2.7188 | 0.0071 | -0.0395 | -0.0063 | TRUE |
| Total.Amygdala \| TotalGrayVol+CAT12 | -0.0229 | 0.0092 | -2.4925 | 0.0135 | -0.0411 | -0.0048 | TRUE |
| Total.Amygdala \| EstimatedTotalIntraCranialVol | -0.0229 | 0.0084 | -2.7359 | 0.0068 | -0.0395 | -0.0064 | TRUE |
| Total.Amygdala \| TotalGrayVol | -0.0230 | 0.0091 | -2.5128 | 0.0128 | -0.0410 | -0.0049 | TRUE |
| Total.Amygdala \| age+sex+BrainSegVol+race+parent_educ+puberty | -0.0230 | 0.0097 | -2.3633 | 0.0192 | -0.0422 | -0.0038 | TRUE |
| Total.Amygdala \| CAT12+race+parent_educ+puberty | -0.0230 | 0.0070 | -3.2848 | 0.0012 | -0.0369 | -0.0092 | TRUE |
| Total.Amygdala \| race+parent_educ+puberty | -0.0231 | 0.0070 | -3.3001 | 0.0012 | -0.0368 | -0.0093 | TRUE |
| Total.Amygdala \| TotalGrayVol+CAT12+parent_educ+puberty | -0.0231 | 0.0095 | -2.4273 | 0.0162 | -0.0418 | -0.0043 | TRUE |
| Total.Amygdala \| age+sex+TotalGrayVol+parent_educ+puberty | -0.0231 | 0.0094 | -2.4446 | 0.0155 | -0.0417 | -0.0045 | TRUE |
| Total.Amygdala \| TotalGrayVol+parent_educ+puberty | -0.0231 | 0.0095 | -2.4442 | 0.0155 | -0.0418 | -0.0045 | TRUE |
| Total.Amygdala \| age+sex+BrainSegVol+CAT12+race | -0.0231 | 0.0095 | -2.4260 | 0.0163 | -0.0420 | -0.0043 | TRUE |
| Total.Amygdala \| BrainSegVol+CAT12+race+parent_educ | -0.0232 | 0.0100 | -2.3218 | 0.0214 | -0.0429 | -0.0035 | TRUE |
| Total.Amygdala \| BrainSegVol+race+parent_educ | -0.0232 | 0.0100 | -2.3304 | 0.0209 | -0.0429 | -0.0036 | TRUE |
| Total.Amygdala \| age+sex+TotalGrayVol | -0.0232 | 0.0091 | -2.5607 | 0.0112 | -0.0411 | -0.0053 | TRUE |
| Total.Amygdala \| TotalGrayVol+race+puberty | -0.0234 | 0.0094 | -2.4899 | 0.0137 | -0.0420 | -0.0049 | TRUE |
| Total.Amygdala \| TotalGrayVol+CAT12+race+puberty | -0.0234 | 0.0094 | -2.4803 | 0.0141 | -0.0420 | -0.0048 | TRUE |
| Total.Amygdala \| age+sex+BrainSegVol+race | -0.0234 | 0.0095 | -2.4601 | 0.0148 | -0.0422 | -0.0046 | TRUE |
| Total.Amygdala \| CAT12+parent_educ | -0.0237 | 0.0070 | -3.3985 | 0.0008 | -0.0375 | -0.0100 | TRUE |
| Total.Amygdala \| EstimatedTotalIntraCranialVol+CAT12+puberty | -0.0238 | 0.0083 | -2.8685 | 0.0046 | -0.0401 | -0.0074 | TRUE |
| Total.Amygdala \| parent_educ | -0.0238 | 0.0070 | -3.4180 | 0.0008 | -0.0375 | -0.0101 | TRUE |
| Total.Amygdala \| EstimatedTotalIntraCranialVol+puberty | -0.0238 | 0.0082 | -2.8891 | 0.0043 | -0.0401 | -0.0076 | TRUE |
| Total.Amygdala \| age+sex+BrainSegVol+CAT12+parent_educ | -0.0238 | 0.0097 | -2.4622 | 0.0147 | -0.0429 | -0.0047 | TRUE |
| Total.Amygdala \| age+sex+BrainSegVol+parent_educ | -0.0240 | 0.0097 | -2.4837 | 0.0139 | -0.0431 | -0.0049 | TRUE |
| Total.Amygdala \| age+sex+TotalGrayVol+CAT12+puberty | -0.0242 | 0.0091 | -2.6768 | 0.0081 | -0.0421 | -0.0064 | TRUE |
| Total.Amygdala \| race | -0.0243 | 0.0067 | -3.6037 | 0.0004 | -0.0376 | -0.0110 | TRUE |
| Total.Amygdala \| CAT12+race | -0.0243 | 0.0068 | -3.5923 | 0.0004 | -0.0377 | -0.0110 | TRUE |
| Total.Amygdala \| age+sex+BrainSegVol+CAT12+race+puberty | -0.0244 | 0.0095 | -2.5691 | 0.0110 | -0.0431 | -0.0057 | TRUE |
| Total.Amygdala \| CAT12+parent_educ+puberty | -0.0245 | 0.0069 | -3.5692 | 0.0005 | -0.0381 | -0.0110 | TRUE |
| Total.Amygdala \| parent_educ+puberty | -0.0246 | 0.0069 | -3.5872 | 0.0004 | -0.0381 | -0.0111 | TRUE |
| Total.Amygdala \| BrainSegVol+CAT12+race+parent_educ+puberty | -0.0246 | 0.0099 | -2.4996 | 0.0134 | -0.0441 | -0.0052 | TRUE |
| Total.Amygdala \| BrainSegVol+race+parent_educ+puberty | -0.0246 | 0.0098 | -2.5085 | 0.0130 | -0.0440 | -0.0053 | TRUE |
| Total.Amygdala \| age+sex+BrainSegVol+race+puberty | -0.0247 | 0.0095 | -2.6038 | 0.0100 | -0.0433 | -0.0060 | TRUE |
| Total.Amygdala \| age+sex+TotalGrayVol+puberty | -0.0247 | 0.0090 | -2.7355 | 0.0068 | -0.0424 | -0.0069 | TRUE |
| Total.Amygdala \| TotalGrayVol+CAT12+puberty | -0.0247 | 0.0091 | -2.7219 | 0.0071 | -0.0426 | -0.0068 | TRUE |
| Total.Amygdala \| TotalGrayVol+puberty | -0.0248 | 0.0090 | -2.7469 | 0.0066 | -0.0426 | -0.0070 | TRUE |
| Total.Amygdala \| BrainSegVol+CAT12+parent_educ | -0.0248 | 0.0098 | -2.5275 | 0.0123 | -0.0442 | -0.0055 | TRUE |
| Total.Amygdala \| BrainSegVol+parent_educ | -0.0249 | 0.0098 | -2.5364 | 0.0120 | -0.0442 | -0.0055 | TRUE |
| Total.Amygdala \| race+puberty | -0.0249 | 0.0066 | -3.7465 | 0.0002 | -0.0380 | -0.0118 | TRUE |
| Total.Amygdala \| CAT12+race+puberty | -0.0249 | 0.0067 | -3.7351 | 0.0003 | -0.0380 | -0.0117 | TRUE |
| Total.Amygdala \| age+sex+BrainSegVol+CAT12+parent_educ+puberty | -0.0253 | 0.0096 | -2.6251 | 0.0094 | -0.0443 | -0.0063 | TRUE |
| Total.Amygdala \| BrainSegVol+race | -0.0253 | 0.0097 | -2.6050 | 0.0100 | -0.0445 | -0.0061 | TRUE |
| Total.Amygdala \| BrainSegVol+CAT12+race | -0.0253 | 0.0098 | -2.5978 | 0.0102 | -0.0446 | -0.0061 | TRUE |
| Total.Amygdala \| age+sex+BrainSegVol+parent_educ+puberty | -0.0255 | 0.0096 | -2.6495 | 0.0088 | -0.0444 | -0.0065 | TRUE |
| Total.Amygdala \| age+sex+BrainSegVol+CAT12 | -0.0255 | 0.0092 | -2.7667 | 0.0062 | -0.0437 | -0.0073 | TRUE |
| Total.Amygdala \| age+sex+BrainSegVol | -0.0258 | 0.0092 | -2.8032 | 0.0056 | -0.0440 | -0.0076 | TRUE |
| Total.Amygdala \| BrainSegVol+CAT12+parent_educ+puberty | -0.0264 | 0.0097 | -2.7249 | 0.0071 | -0.0455 | -0.0073 | TRUE |
| Total.Amygdala \| CAT12 | -0.0264 | 0.0065 | -4.0755 | 0.0001 | -0.0392 | -0.0136 | TRUE |
| Total.Amygdala \| BrainSegVol+parent_educ+puberty | -0.0264 | 0.0097 | -2.7344 | 0.0069 | -0.0455 | -0.0074 | TRUE |
| Total.Amygdala \| unadjusted | -0.0265 | 0.0064 | -4.1043 | 0.0001 | -0.0392 | -0.0138 | TRUE |
| Total.Amygdala \| BrainSegVol+race+puberty | -0.0266 | 0.0096 | -2.7753 | 0.0061 | -0.0455 | -0.0077 | TRUE |
| Total.Amygdala \| BrainSegVol+CAT12+race+puberty | -0.0266 | 0.0096 | -2.7677 | 0.0062 | -0.0455 | -0.0076 | TRUE |
| Total.Amygdala \| BrainSegVol+CAT12 | -0.0266 | 0.0094 | -2.8429 | 0.0050 | -0.0451 | -0.0081 | TRUE |
| Total.Amygdala \| BrainSegVol | -0.0266 | 0.0093 | -2.8544 | 0.0048 | -0.0450 | -0.0082 | TRUE |
| Total.Amygdala \| CAT12+puberty | -0.0269 | 0.0064 | -4.2179 | 0.0000 | -0.0395 | -0.0143 | TRUE |
| Total.Amygdala \| puberty | -0.0270 | 0.0063 | -4.2506 | 0.0000 | -0.0395 | -0.0145 | TRUE |
| Total.Amygdala \| age+sex+BrainSegVol+CAT12+puberty | -0.0270 | 0.0092 | -2.9465 | 0.0036 | -0.0451 | -0.0089 | TRUE |
| Total.Amygdala \| age+sex+BrainSegVol+puberty | -0.0273 | 0.0091 | -2.9840 | 0.0032 | -0.0453 | -0.0093 | TRUE |
| Total.Amygdala \| BrainSegVol+CAT12+puberty | -0.0282 | 0.0092 | -3.0619 | 0.0025 | -0.0464 | -0.0100 | TRUE |
| Total.Amygdala \| BrainSegVol+puberty | -0.0283 | 0.0092 | -3.0758 | 0.0024 | -0.0464 | -0.0101 | TRUE |
| Right.Amygdala \| age+sex+CAT12+puberty | -0.0289 | 0.0140 | -2.0712 | 0.0397 | -0.0565 | -0.0014 | TRUE |
| Right.Amygdala \| age+sex+puberty | -0.0303 | 0.0138 | -2.1909 | 0.0297 | -0.0575 | -0.0030 | TRUE |
| Right.Amygdala \| EstimatedTotalIntraCranialVol | -0.0310 | 0.0156 | -1.9816 | 0.0489 | -0.0618 | -0.0001 | TRUE |
| Left.Amygdala \| age+sex+CAT12+race+parent_educ | -0.0312 | 0.0148 | -2.1055 | 0.0367 | -0.0604 | -0.0020 | TRUE |
| Left.Amygdala \| age+sex+race+parent_educ | -0.0322 | 0.0147 | -2.1922 | 0.0297 | -0.0612 | -0.0032 | TRUE |
| Right.Amygdala \| EstimatedTotalIntraCranialVol+CAT12+puberty | -0.0323 | 0.0155 | -2.0886 | 0.0381 | -0.0628 | -0.0018 | TRUE |
| Right.Amygdala \| EstimatedTotalIntraCranialVol+puberty | -0.0324 | 0.0154 | -2.1046 | 0.0366 | -0.0628 | -0.0020 | TRUE |
| Left.Amygdala \| age+sex+CAT12+race | -0.0326 | 0.0142 | -2.3035 | 0.0224 | -0.0606 | -0.0047 | TRUE |
| Right.Amygdala \| age+sex+BrainSegVol+CAT12+puberty | -0.0333 | 0.0169 | -1.9773 | 0.0495 | -0.0666 | -0.0001 | TRUE |
| Right.Amygdala \| CAT12+race+parent_educ | -0.0336 | 0.0133 | -2.5323 | 0.0122 | -0.0597 | -0.0074 | TRUE |
| Right.Amygdala \| race+parent_educ | -0.0336 | 0.0132 | -2.5472 | 0.0117 | -0.0597 | -0.0076 | TRUE |
| Left.Amygdala \| age+sex+race | -0.0337 | 0.0140 | -2.3984 | 0.0175 | -0.0614 | -0.0060 | TRUE |
| Right.Amygdala \| age+sex+BrainSegVol+puberty | -0.0338 | 0.0168 | -2.0081 | 0.0461 | -0.0670 | -0.0006 | TRUE |
| Right.Amygdala \| BrainSegVol+CAT12 | -0.0345 | 0.0171 | -2.0162 | 0.0452 | -0.0683 | -0.0008 | TRUE |
| Right.Amygdala \| BrainSegVol | -0.0345 | 0.0171 | -2.0232 | 0.0444 | -0.0682 | -0.0009 | TRUE |
| Left.Amygdala \| age+sex+CAT12+race+parent_educ+puberty | -0.0349 | 0.0148 | -2.3606 | 0.0194 | -0.0641 | -0.0057 | TRUE |
| Right.Amygdala \| BrainSegVol+race+puberty | -0.0350 | 0.0177 | -1.9777 | 0.0495 | -0.0699 | -0.0001 | TRUE |
| Right.Amygdala \| CAT12+race+parent_educ+puberty | -0.0354 | 0.0131 | -2.7073 | 0.0075 | -0.0612 | -0.0096 | TRUE |
| Right.Amygdala \| race+parent_educ+puberty | -0.0354 | 0.0130 | -2.7213 | 0.0072 | -0.0611 | -0.0097 | TRUE |
| Left.Amygdala \| age+sex+race+parent_educ+puberty | -0.0358 | 0.0147 | -2.4410 | 0.0156 | -0.0648 | -0.0069 | TRUE |
| Left.Amygdala \| age+sex+EstimatedTotalIntraCranialVol+CAT12+race+parent_educ | -0.0359 | 0.0160 | -2.2405 | 0.0263 | -0.0676 | -0.0043 | TRUE |
| Left.Amygdala \| age+sex+CAT12+parent_educ | -0.0361 | 0.0145 | -2.4836 | 0.0139 | -0.0647 | -0.0074 | TRUE |
| Left.Amygdala \| age+sex+CAT12+race+puberty | -0.0361 | 0.0141 | -2.5522 | 0.0116 | -0.0640 | -0.0082 | TRUE |
| Left.Amygdala \| age+sex+EstimatedTotalIntraCranialVol+race+parent_educ | -0.0366 | 0.0160 | -2.2891 | 0.0233 | -0.0682 | -0.0050 | TRUE |
| Right.Amygdala \| CAT12+parent_educ | -0.0369 | 0.0130 | -2.8455 | 0.0049 | -0.0625 | -0.0113 | TRUE |
| Left.Amygdala \| age+sex+race+puberty | -0.0370 | 0.0140 | -2.6393 | 0.0090 | -0.0646 | -0.0093 | TRUE |
| Right.Amygdala \| parent_educ | -0.0370 | 0.0129 | -2.8639 | 0.0047 | -0.0625 | -0.0115 | TRUE |
| Left.Amygdala \| age+sex+parent_educ | -0.0370 | 0.0144 | -2.5637 | 0.0112 | -0.0655 | -0.0085 | TRUE |
| Right.Amygdala \| BrainSegVol+CAT12+puberty | -0.0371 | 0.0169 | -2.1948 | 0.0294 | -0.0704 | -0.0038 | TRUE |
| Right.Amygdala \| BrainSegVol+puberty | -0.0371 | 0.0169 | -2.2030 | 0.0288 | -0.0704 | -0.0039 | TRUE |
| Left.Amygdala \| age+sex+EstimatedTotalIntraCranialVol+CAT12+race | -0.0375 | 0.0155 | -2.4195 | 0.0166 | -0.0681 | -0.0069 | TRUE |
| Left.Amygdala \| age+sex+EstimatedTotalIntraCranialVol+CAT12+race+parent_educ+puberty | -0.0380 | 0.0160 | -2.3831 | 0.0183 | -0.0695 | -0.0065 | TRUE |
| Left.Amygdala \| age+sex+EstimatedTotalIntraCranialVol+race | -0.0382 | 0.0154 | -2.4736 | 0.0143 | -0.0687 | -0.0077 | TRUE |
| Right.Amygdala \| CAT12+parent_educ+puberty | -0.0382 | 0.0128 | -2.9924 | 0.0031 | -0.0635 | -0.0130 | TRUE |
| Right.Amygdala \| parent_educ+puberty | -0.0383 | 0.0127 | -3.0094 | 0.0030 | -0.0634 | -0.0132 | TRUE |
| Left.Amygdala \| age+sex+EstimatedTotalIntraCranialVol+race+parent_educ+puberty | -0.0386 | 0.0159 | -2.4281 | 0.0162 | -0.0700 | -0.0072 | TRUE |
| Right.Amygdala \| CAT12+race | -0.0386 | 0.0128 | -3.0193 | 0.0029 | -0.0639 | -0.0134 | TRUE |
| Right.Amygdala \| race | -0.0386 | 0.0127 | -3.0294 | 0.0028 | -0.0638 | -0.0135 | TRUE |
| Left.Amygdala \| age+sex+EstimatedTotalIntraCranialVol+CAT12+race+puberty | -0.0393 | 0.0154 | -2.5519 | 0.0116 | -0.0697 | -0.0089 | TRUE |
| Left.Amygdala \| age+sex+CAT12+parent_educ+puberty | -0.0393 | 0.0145 | -2.7188 | 0.0072 | -0.0679 | -0.0108 | TRUE |
| Left.Amygdala \| age+sex+EstimatedTotalIntraCranialVol+CAT12+parent_educ | -0.0396 | 0.0158 | -2.5010 | 0.0133 | -0.0709 | -0.0084 | TRUE |
| Right.Amygdala \| race+puberty | -0.0397 | 0.0125 | -3.1635 | 0.0018 | -0.0644 | -0.0149 | TRUE |
| Right.Amygdala \| CAT12+race+puberty | -0.0397 | 0.0126 | -3.1534 | 0.0019 | -0.0645 | -0.0149 | TRUE |
| Left.Amygdala \| age+sex+CAT12 | -0.0397 | 0.0136 | -2.9210 | 0.0039 | -0.0666 | -0.0129 | TRUE |
| Left.Amygdala \| age+sex+EstimatedTotalIntraCranialVol+race+puberty | -0.0399 | 0.0153 | -2.6007 | 0.0101 | -0.0702 | -0.0096 | TRUE |
| Left.Amygdala \| EstimatedTotalIntraCranialVol+CAT12+race+parent_educ | -0.0399 | 0.0162 | -2.4649 | 0.0147 | -0.0719 | -0.0080 | TRUE |
| Left.Amygdala \| EstimatedTotalIntraCranialVol+race+parent_educ | -0.0400 | 0.0161 | -2.4765 | 0.0142 | -0.0718 | -0.0081 | TRUE |
| Left.Amygdala \| age+sex+parent_educ+puberty | -0.0401 | 0.0144 | -2.7919 | 0.0058 | -0.0685 | -0.0118 | TRUE |
| Left.Amygdala \| age+sex+EstimatedTotalIntraCranialVol+parent_educ | -0.0402 | 0.0158 | -2.5390 | 0.0120 | -0.0714 | -0.0089 | TRUE |
| Left.Amygdala \| TotalGrayVol+CAT12+race+parent_educ | -0.0406 | 0.0173 | -2.3464 | 0.0201 | -0.0747 | -0.0064 | TRUE |
| Left.Amygdala \| TotalGrayVol+race+parent_educ | -0.0407 | 0.0172 | -2.3611 | 0.0193 | -0.0746 | -0.0067 | TRUE |
| Left.Amygdala \| age+sex | -0.0409 | 0.0135 | -3.0411 | 0.0027 | -0.0675 | -0.0144 | TRUE |
| Left.Amygdala \| age+sex+EstimatedTotalIntraCranialVol+CAT12+parent_educ+puberty | -0.0414 | 0.0157 | -2.6312 | 0.0092 | -0.0724 | -0.0104 | TRUE |
| Left.Amygdala \| EstimatedTotalIntraCranialVol+CAT12+race+parent_educ+puberty | -0.0417 | 0.0159 | -2.6135 | 0.0097 | -0.0732 | -0.0102 | TRUE |
| Left.Amygdala \| age+sex+TotalGrayVol+CAT12+race+parent_educ | -0.0417 | 0.0170 | -2.4563 | 0.0150 | -0.0752 | -0.0082 | TRUE |
| Left.Amygdala \| EstimatedTotalIntraCranialVol+race+parent_educ+puberty | -0.0417 | 0.0159 | -2.6246 | 0.0094 | -0.0731 | -0.0103 | TRUE |
| Left.Amygdala \| EstimatedTotalIntraCranialVol+CAT12+parent_educ | -0.0418 | 0.0159 | -2.6218 | 0.0095 | -0.0733 | -0.0103 | TRUE |
| Left.Amygdala \| age+sex+EstimatedTotalIntraCranialVol+parent_educ+puberty | -0.0418 | 0.0157 | -2.6660 | 0.0084 | -0.0728 | -0.0109 | TRUE |
| Left.Amygdala \| EstimatedTotalIntraCranialVol+parent_educ | -0.0419 | 0.0159 | -2.6346 | 0.0091 | -0.0733 | -0.0105 | TRUE |
| Left.Amygdala \| TotalGrayVol+CAT12+parent_educ | -0.0420 | 0.0170 | -2.4738 | 0.0143 | -0.0755 | -0.0085 | TRUE |
| Left.Amygdala \| TotalGrayVol+parent_educ | -0.0421 | 0.0169 | -2.4901 | 0.0137 | -0.0755 | -0.0088 | TRUE |
| Right.Amygdala \| CAT12 | -0.0423 | 0.0122 | -3.4574 | 0.0007 | -0.0664 | -0.0182 | TRUE |
| Left.Amygdala \| age+sex+TotalGrayVol+race+parent_educ | -0.0423 | 0.0169 | -2.4987 | 0.0134 | -0.0757 | -0.0089 | TRUE |
| Right.Amygdala \| unadjusted | -0.0424 | 0.0122 | -3.4835 | 0.0006 | -0.0664 | -0.0184 | TRUE |
| Left.Amygdala \| age+sex+EstimatedTotalIntraCranialVol+CAT12 | -0.0424 | 0.0150 | -2.8221 | 0.0053 | -0.0721 | -0.0128 | TRUE |
| Left.Amygdala \| EstimatedTotalIntraCranialVol+race | -0.0427 | 0.0156 | -2.7337 | 0.0069 | -0.0736 | -0.0119 | TRUE |
| Left.Amygdala \| EstimatedTotalIntraCranialVol+CAT12+race | -0.0427 | 0.0157 | -2.7251 | 0.0071 | -0.0737 | -0.0118 | TRUE |
| Left.Amygdala \| age+sex+CAT12+puberty | -0.0430 | 0.0136 | -3.1745 | 0.0018 | -0.0698 | -0.0163 | TRUE |
| Right.Amygdala \| CAT12+puberty | -0.0430 | 0.0120 | -3.5765 | 0.0004 | -0.0668 | -0.0193 | TRUE |
| Left.Amygdala \| TotalGrayVol+CAT12+race+parent_educ+puberty | -0.0432 | 0.0170 | -2.5336 | 0.0122 | -0.0768 | -0.0095 | TRUE |
| Right.Amygdala \| puberty | -0.0432 | 0.0120 | -3.6056 | 0.0004 | -0.0669 | -0.0196 | TRUE |
| Left.Amygdala \| TotalGrayVol+race+parent_educ+puberty | -0.0432 | 0.0170 | -2.5476 | 0.0117 | -0.0767 | -0.0097 | TRUE |
| Left.Amygdala \| age+sex+EstimatedTotalIntraCranialVol | -0.0432 | 0.0150 | -2.8891 | 0.0043 | -0.0728 | -0.0137 | TRUE |
| Left.Amygdala \| TotalGrayVol+race | -0.0433 | 0.0168 | -2.5783 | 0.0107 | -0.0765 | -0.0102 | TRUE |
| Left.Amygdala \| TotalGrayVol+CAT12+race | -0.0434 | 0.0169 | -2.5685 | 0.0110 | -0.0767 | -0.0100 | TRUE |
| Left.Amygdala \| EstimatedTotalIntraCranialVol+CAT12+parent_educ+puberty | -0.0434 | 0.0157 | -2.7643 | 0.0063 | -0.0744 | -0.0124 | TRUE |
| Left.Amygdala \| age+sex+TotalGrayVol+CAT12+race | -0.0434 | 0.0165 | -2.6321 | 0.0092 | -0.0760 | -0.0109 | TRUE |
| Left.Amygdala \| age+sex+TotalGrayVol+CAT12+race+parent_educ+puberty | -0.0434 | 0.0169 | -2.5742 | 0.0109 | -0.0768 | -0.0101 | TRUE |
| Left.Amygdala \| EstimatedTotalIntraCranialVol+parent_educ+puberty | -0.0435 | 0.0157 | -2.7764 | 0.0061 | -0.0743 | -0.0126 | TRUE |
| Left.Amygdala \| EstimatedTotalIntraCranialVol+race+puberty | -0.0438 | 0.0154 | -2.8509 | 0.0049 | -0.0742 | -0.0135 | TRUE |
| Left.Amygdala \| EstimatedTotalIntraCranialVol+CAT12+race+puberty | -0.0438 | 0.0154 | -2.8421 | 0.0050 | -0.0743 | -0.0134 | TRUE |
| Left.Amygdala \| age+sex+TotalGrayVol+race+parent_educ+puberty | -0.0440 | 0.0168 | -2.6145 | 0.0097 | -0.0772 | -0.0108 | TRUE |
| Left.Amygdala \| age+sex+TotalGrayVol+race | -0.0441 | 0.0165 | -2.6772 | 0.0081 | -0.0765 | -0.0116 | TRUE |
| Left.Amygdala \| age+sex+puberty | -0.0441 | 0.0134 | -3.2900 | 0.0012 | -0.0705 | -0.0177 | TRUE |
| Left.Amygdala \| age+sex+TotalGrayVol+CAT12+parent_educ | -0.0442 | 0.0168 | -2.6317 | 0.0092 | -0.0774 | -0.0111 | TRUE |
| Left.Amygdala \| age+sex+EstimatedTotalIntraCranialVol+CAT12+puberty | -0.0443 | 0.0149 | -2.9690 | 0.0034 | -0.0738 | -0.0149 | TRUE |
| Left.Amygdala \| age+sex+TotalGrayVol+parent_educ | -0.0447 | 0.0168 | -2.6631 | 0.0084 | -0.0778 | -0.0116 | TRUE |
| Left.Amygdala \| TotalGrayVol+CAT12+parent_educ+puberty | -0.0448 | 0.0167 | -2.6748 | 0.0082 | -0.0778 | -0.0118 | TRUE |
| Left.Amygdala \| TotalGrayVol+parent_educ+puberty | -0.0449 | 0.0167 | -2.6906 | 0.0078 | -0.0778 | -0.0120 | TRUE |
| Left.Amygdala \| age+sex+TotalGrayVol+CAT12+race+puberty | -0.0449 | 0.0164 | -2.7377 | 0.0068 | -0.0773 | -0.0125 | TRUE |
| Left.Amygdala \| TotalGrayVol+CAT12 | -0.0449 | 0.0162 | -2.7653 | 0.0062 | -0.0769 | -0.0129 | TRUE |
| Left.Amygdala \| CAT12+race+parent_educ | -0.0449 | 0.0136 | -3.2919 | 0.0012 | -0.0718 | -0.0180 | TRUE |
| Left.Amygdala \| race+parent_educ | -0.0450 | 0.0136 | -3.3080 | 0.0011 | -0.0718 | -0.0181 | TRUE |
| Left.Amygdala \| TotalGrayVol | -0.0450 | 0.0162 | -2.7864 | 0.0059 | -0.0769 | -0.0131 | TRUE |
| Left.Amygdala \| age+sex+EstimatedTotalIntraCranialVol+puberty | -0.0450 | 0.0149 | -3.0319 | 0.0028 | -0.0743 | -0.0157 | TRUE |
| Left.Amygdala \| EstimatedTotalIntraCranialVol+CAT12 | -0.0451 | 0.0151 | -2.9797 | 0.0033 | -0.0750 | -0.0153 | TRUE |
| Left.Amygdala \| age+sex+BrainSegVol+CAT12+race+parent_educ | -0.0452 | 0.0172 | -2.6225 | 0.0095 | -0.0792 | -0.0112 | TRUE |
| Left.Amygdala \| EstimatedTotalIntraCranialVol | -0.0452 | 0.0151 | -2.9977 | 0.0031 | -0.0749 | -0.0155 | TRUE |
| Left.Amygdala \| TotalGrayVol+race+puberty | -0.0454 | 0.0165 | -2.7426 | 0.0067 | -0.0780 | -0.0127 | TRUE |
| Left.Amygdala \| TotalGrayVol+CAT12+race+puberty | -0.0454 | 0.0166 | -2.7325 | 0.0069 | -0.0782 | -0.0126 | TRUE |
| Left.Amygdala \| age+sex+TotalGrayVol+race+puberty | -0.0454 | 0.0163 | -2.7795 | 0.0060 | -0.0777 | -0.0132 | TRUE |
| Left.Amygdala \| age+sex+BrainSegVol+race+parent_educ | -0.0455 | 0.0172 | -2.6413 | 0.0090 | -0.0794 | -0.0115 | TRUE |
| Left.Amygdala \| BrainSegVol+CAT12+race+parent_educ | -0.0455 | 0.0177 | -2.5752 | 0.0108 | -0.0803 | -0.0106 | TRUE |
| Left.Amygdala \| BrainSegVol+race+parent_educ | -0.0455 | 0.0176 | -2.5842 | 0.0106 | -0.0802 | -0.0108 | TRUE |
| Left.Amygdala \| age+sex+TotalGrayVol+CAT12+parent_educ+puberty | -0.0461 | 0.0167 | -2.7619 | 0.0063 | -0.0790 | -0.0132 | TRUE |
| Left.Amygdala \| age+sex+TotalGrayVol+CAT12 | -0.0464 | 0.0160 | -2.8903 | 0.0043 | -0.0780 | -0.0147 | TRUE |
| Left.Amygdala \| age+sex+TotalGrayVol+parent_educ+puberty | -0.0465 | 0.0166 | -2.7925 | 0.0058 | -0.0793 | -0.0136 | TRUE |
| Left.Amygdala \| EstimatedTotalIntraCranialVol+CAT12+puberty | -0.0466 | 0.0149 | -3.1316 | 0.0020 | -0.0760 | -0.0173 | TRUE |
| Left.Amygdala \| EstimatedTotalIntraCranialVol+puberty | -0.0468 | 0.0148 | -3.1536 | 0.0019 | -0.0760 | -0.0175 | TRUE |
| Left.Amygdala \| age+sex+BrainSegVol+CAT12+race+parent_educ+puberty | -0.0469 | 0.0171 | -2.7354 | 0.0069 | -0.0807 | -0.0130 | TRUE |
| Left.Amygdala \| age+sex+BrainSegVol+CAT12+race | -0.0469 | 0.0167 | -2.8084 | 0.0055 | -0.0798 | -0.0139 | TRUE |
| Left.Amygdala \| CAT12+race+parent_educ+puberty | -0.0469 | 0.0135 | -3.4875 | 0.0006 | -0.0735 | -0.0204 | TRUE |
| Left.Amygdala \| race+parent_educ+puberty | -0.0470 | 0.0134 | -3.5028 | 0.0006 | -0.0734 | -0.0205 | TRUE |
| Left.Amygdala \| age+sex+TotalGrayVol | -0.0471 | 0.0160 | -2.9483 | 0.0036 | -0.0786 | -0.0156 | TRUE |
| Left.Amygdala \| age+sex+BrainSegVol+race+parent_educ+puberty | -0.0471 | 0.0171 | -2.7561 | 0.0065 | -0.0809 | -0.0134 | TRUE |
| Left.Amygdala \| age+sex+BrainSegVol+race | -0.0472 | 0.0167 | -2.8325 | 0.0052 | -0.0801 | -0.0143 | TRUE |
| Left.Amygdala \| BrainSegVol+CAT12+parent_educ | -0.0473 | 0.0174 | -2.7238 | 0.0071 | -0.0816 | -0.0130 | TRUE |
| Left.Amygdala \| BrainSegVol+parent_educ | -0.0474 | 0.0173 | -2.7326 | 0.0069 | -0.0815 | -0.0132 | TRUE |
| Left.Amygdala \| CAT12+parent_educ | -0.0477 | 0.0134 | -3.5554 | 0.0005 | -0.0742 | -0.0212 | TRUE |
| Left.Amygdala \| TotalGrayVol+CAT12+puberty | -0.0477 | 0.0160 | -2.9826 | 0.0032 | -0.0793 | -0.0162 | TRUE |
| Left.Amygdala \| BrainSegVol+CAT12+race+parent_educ+puberty | -0.0477 | 0.0174 | -2.7441 | 0.0067 | -0.0821 | -0.0134 | TRUE |
| Left.Amygdala \| BrainSegVol+race+parent_educ+puberty | -0.0477 | 0.0173 | -2.7534 | 0.0065 | -0.0820 | -0.0135 | TRUE |
| Left.Amygdala \| parent_educ | -0.0478 | 0.0134 | -3.5743 | 0.0004 | -0.0742 | -0.0214 | TRUE |
| Left.Amygdala \| TotalGrayVol+puberty | -0.0479 | 0.0159 | -3.0084 | 0.0030 | -0.0793 | -0.0165 | TRUE |
| Left.Amygdala \| age+sex+TotalGrayVol+CAT12+puberty | -0.0483 | 0.0159 | -3.0351 | 0.0027 | -0.0797 | -0.0169 | TRUE |
| Left.Amygdala \| age+sex+BrainSegVol+CAT12+race+puberty | -0.0483 | 0.0166 | -2.9123 | 0.0041 | -0.0811 | -0.0156 | TRUE |
| Left.Amygdala \| age+sex+BrainSegVol+CAT12+parent_educ | -0.0484 | 0.0171 | -2.8384 | 0.0050 | -0.0821 | -0.0148 | TRUE |
| Left.Amygdala \| BrainSegVol+race | -0.0484 | 0.0171 | -2.8292 | 0.0052 | -0.0822 | -0.0147 | TRUE |
| Left.Amygdala \| BrainSegVol+CAT12+race | -0.0484 | 0.0172 | -2.8213 | 0.0053 | -0.0823 | -0.0146 | TRUE |
| Left.Amygdala \| age+sex+BrainSegVol+parent_educ | -0.0485 | 0.0171 | -2.8465 | 0.0049 | -0.0822 | -0.0149 | TRUE |
| Left.Amygdala \| race | -0.0486 | 0.0129 | -3.7783 | 0.0002 | -0.0739 | -0.0232 | TRUE |
| Left.Amygdala \| CAT12+race | -0.0486 | 0.0129 | -3.7665 | 0.0002 | -0.0740 | -0.0231 | TRUE |
| Left.Amygdala \| age+sex+BrainSegVol+race+puberty | -0.0487 | 0.0166 | -2.9369 | 0.0038 | -0.0814 | -0.0160 | TRUE |
| Left.Amygdala \| age+sex+TotalGrayVol+puberty | -0.0490 | 0.0159 | -3.0908 | 0.0023 | -0.0803 | -0.0177 | TRUE |
| Left.Amygdala \| CAT12+parent_educ+puberty | -0.0493 | 0.0132 | -3.7283 | 0.0003 | -0.0754 | -0.0232 | TRUE |
| Left.Amygdala \| parent_educ+puberty | -0.0494 | 0.0132 | -3.7457 | 0.0002 | -0.0754 | -0.0234 | TRUE |
| Left.Amygdala \| race+puberty | -0.0495 | 0.0127 | -3.9124 | 0.0001 | -0.0745 | -0.0245 | TRUE |
| Left.Amygdala \| CAT12+race+puberty | -0.0495 | 0.0127 | -3.9006 | 0.0001 | -0.0746 | -0.0245 | TRUE |
| Left.Amygdala \| BrainSegVol+CAT12+parent_educ+puberty | -0.0498 | 0.0171 | -2.9106 | 0.0041 | -0.0836 | -0.0161 | TRUE |
| Left.Amygdala \| BrainSegVol+parent_educ+puberty | -0.0499 | 0.0171 | -2.9200 | 0.0039 | -0.0835 | -0.0162 | TRUE |
| Left.Amygdala \| BrainSegVol+race+puberty | -0.0501 | 0.0168 | -2.9771 | 0.0033 | -0.0833 | -0.0169 | TRUE |
| Left.Amygdala \| BrainSegVol+CAT12+race+puberty | -0.0501 | 0.0169 | -2.9689 | 0.0034 | -0.0835 | -0.0168 | TRUE |
| Left.Amygdala \| age+sex+BrainSegVol+CAT12+parent_educ+puberty | -0.0502 | 0.0169 | -2.9644 | 0.0034 | -0.0836 | -0.0168 | TRUE |
| Left.Amygdala \| age+sex+BrainSegVol+parent_educ+puberty | -0.0503 | 0.0169 | -2.9758 | 0.0033 | -0.0837 | -0.0170 | TRUE |
| Left.Amygdala \| BrainSegVol+CAT12 | -0.0505 | 0.0165 | -3.0548 | 0.0026 | -0.0831 | -0.0179 | TRUE |
| Left.Amygdala \| BrainSegVol | -0.0506 | 0.0165 | -3.0677 | 0.0025 | -0.0831 | -0.0181 | TRUE |
| Left.Amygdala \| age+sex+BrainSegVol+CAT12 | -0.0509 | 0.0162 | -3.1329 | 0.0020 | -0.0829 | -0.0188 | TRUE |
| Left.Amygdala \| age+sex+BrainSegVol | -0.0514 | 0.0162 | -3.1696 | 0.0018 | -0.0833 | -0.0194 | TRUE |
| Left.Amygdala \| CAT12 | -0.0524 | 0.0124 | -4.2293 | 0.0000 | -0.0768 | -0.0280 | TRUE |
| Left.Amygdala \| unadjusted | -0.0524 | 0.0123 | -4.2588 | 0.0000 | -0.0767 | -0.0282 | TRUE |
| Left.Amygdala \| age+sex+BrainSegVol+CAT12+puberty | -0.0528 | 0.0161 | -3.2775 | 0.0012 | -0.0846 | -0.0210 | TRUE |
| Left.Amygdala \| BrainSegVol+CAT12+puberty | -0.0531 | 0.0163 | -3.2625 | 0.0013 | -0.0852 | -0.0210 | TRUE |
| Left.Amygdala \| BrainSegVol+puberty | -0.0532 | 0.0162 | -3.2784 | 0.0012 | -0.0852 | -0.0212 | TRUE |
| Left.Amygdala \| age+sex+BrainSegVol+puberty | -0.0533 | 0.0161 | -3.3141 | 0.0011 | -0.0850 | -0.0216 | TRUE |
| Left.Amygdala \| CAT12+puberty | -0.0534 | 0.0122 | -4.3775 | 0.0000 | -0.0774 | -0.0293 | TRUE |
| Left.Amygdala \| puberty | -0.0535 | 0.0121 | -4.4112 | 0.0000 | -0.0774 | -0.0296 | TRUE |

**Table S3. Multiverse results: hippocampal volume across alternative STRAIN stress outcomes**

*18 model specifications (3 ROI operationalizations × 6 STRAIN stress outcomes). Age, sex, eTIV, and pubertal status were held constant across all specifications. Values rounded to four decimal places.*

| **ROI** | **Stress outcome** | **estimate** | **SE** | **t** | **p** | **CI low** | **CI high** | **significant** |
| --- | --- | --- | --- | --- | --- | --- | --- | --- |
| Total.Hippocampus | Acute count | -0.0002 | 0.0001 | -1.7525 | 0.0813 | -0.0004 | 0.0000 | False |
| Left.Hippocampus | Acute count | -0.0002 | 0.0002 | -0.9907 | 0.3231 | -0.0006 | 0.0002 | False |
| Left.Hippocampus | Total count | -0.0003 | 0.0002 | -1.5906 | 0.1134 | -0.0007 | 0.0001 | False |
| Left.Hippocampus | Acute severity | -0.0004 | 0.0002 | -1.7041 | 0.0900 | -0.0008 | 0.0001 | False |
| Total.Hippocampus | Chronic severity | -0.0002 | 0.0001 | -2.3659 | 0.0190 | -0.0004 | -0.0000 | True |
| Total.Hippocampus | Total count | -0.0002 | 0.0001 | -2.3202 | 0.0214 | -0.0004 | -0.0000 | True |
| Total.Hippocampus | Acute severity | -0.0002 | 0.0001 | -2.3599 | 0.0193 | -0.0005 | -0.0000 | True |
| Total.Hippocampus | Total severity | -0.0003 | 0.0001 | -2.4972 | 0.0134 | -0.0005 | -0.0001 | True |
| Total.Hippocampus | Chronic count | -0.0003 | 0.0001 | -2.6815 | 0.0080 | -0.0005 | -0.0001 | True |
| Left.Hippocampus | Chronic severity | -0.0004 | 0.0002 | -2.0194 | 0.0449 | -0.0008 | -0.0000 | True |
| Left.Hippocampus | Total severity | -0.0004 | 0.0002 | -2.0155 | 0.0453 | -0.0008 | -0.0000 | True |
| Right.Hippocampus | Acute count | -0.0004 | 0.0002 | -2.2413 | 0.0262 | -0.0008 | -0.0001 | True |
| Left.Hippocampus | Chronic count | -0.0004 | 0.0002 | -2.1309 | 0.0344 | -0.0008 | -0.0000 | True |
| Right.Hippocampus | Chronic severity | -0.0004 | 0.0002 | -2.4003 | 0.0173 | -0.0008 | -0.0001 | True |
| Right.Hippocampus | Total severity | -0.0005 | 0.0002 | -2.6386 | 0.0090 | -0.0008 | -0.0001 | True |
| Right.Hippocampus | Acute severity | -0.0005 | 0.0002 | -2.6785 | 0.0080 | -0.0009 | -0.0001 | True |
| Right.Hippocampus | Total count | -0.0005 | 0.0002 | -2.7121 | 0.0073 | -0.0009 | -0.0001 | True |
| Right.Hippocampus | Chronic count | -0.0005 | 0.0002 | -2.8630 | 0.0047 | -0.0009 | -0.0002 | True |

**Table S4. Multiverse results: amygdala volume across alternative STRAIN stress outcomes**

*18 model specifications (3 ROI operationalizations × 6 STRAIN stress outcomes). Age, sex, eTIV, and pubertal status were held constant across all specifications. Values rounded to four decimal places.*

| **ROI** | **Stress outcome** | **estimate** | **SE** | **t** | **p** | **CI low** | **CI high** | **significant** |
| --- | --- | --- | --- | --- | --- | --- | --- | --- |
| Right.Amygdala | Chronic severity | -0.0006 | 0.0004 | -1.3698 | 0.1724 | -0.0014 | 0.0003 | False |
| Right.Amygdala | Chronic count | -0.0007 | 0.0004 | -1.7102 | 0.0889 | -0.0016 | 0.0001 | False |
| Right.Amygdala | Total severity | -0.0008 | 0.0004 | -1.8167 | 0.0708 | -0.0016 | 0.0001 | False |
| Total.Amygdala | Chronic severity | -0.0005 | 0.0002 | -2.1986 | 0.0291 | -0.0009 | -0.0001 | True |
| Total.Amygdala | Total severity | -0.0006 | 0.0002 | -2.6692 | 0.0083 | -0.0010 | -0.0002 | True |
| Total.Amygdala | Chronic count | -0.0006 | 0.0002 | -2.6991 | 0.0076 | -0.0010 | -0.0002 | True |
| Total.Amygdala | Acute severity | -0.0007 | 0.0002 | -3.1478 | 0.0019 | -0.0012 | -0.0003 | True |
| Total.Amygdala | Total count | -0.0007 | 0.0002 | -3.2412 | 0.0014 | -0.0012 | -0.0003 | True |
| Total.Amygdala | Acute count | -0.0007 | 0.0002 | -3.2037 | 0.0016 | -0.0012 | -0.0003 | True |
| Right.Amygdala | Total count | -0.0010 | 0.0004 | -2.2606 | 0.0249 | -0.0018 | -0.0001 | True |
| Right.Amygdala | Acute severity | -0.0010 | 0.0004 | -2.3851 | 0.0181 | -0.0019 | -0.0002 | True |
| Right.Amygdala | Acute count | -0.0010 | 0.0004 | -2.3701 | 0.0188 | -0.0019 | -0.0002 | True |
| Left.Amygdala | Chronic severity | -0.0011 | 0.0004 | -2.6247 | 0.0094 | -0.0018 | -0.0003 | True |
| Left.Amygdala | Total severity | -0.0012 | 0.0004 | -3.0319 | 0.0028 | -0.0020 | -0.0004 | True |
| Left.Amygdala | Chronic count | -0.0013 | 0.0004 | -3.1949 | 0.0016 | -0.0021 | -0.0005 | True |
| Left.Amygdala | Acute severity | -0.0014 | 0.0004 | -3.3276 | 0.0011 | -0.0022 | -0.0006 | True |
| Left.Amygdala | Acute count | -0.0014 | 0.0004 | -3.4442 | 0.0007 | -0.0022 | -0.0006 | True |
| Left.Amygdala | Total count | -0.0015 | 0.0004 | -3.6242 | 0.0004 | -0.0023 | -0.0007 | True |
